# Interictal Mini-Seizures: Recurrent Neuronal Synchronization Events Driven by the Epileptogenic Zone

**DOI:** 10.1101/2025.01.31.25321482

**Authors:** Tonmoy Monsoor, Sotaro Kanai, Prateik Sinha, Atsuro Daida, Naoto Kuroda, Shingo Oana, Lina Zhang, Lawrence Liu, Gaurav Singh, Yipeng Zhang, Chenda Duan, Shaun A. Hussain, Raman Sankar, Aria Fallah, Myung Shin Sim, William Speier, Richard J Staba, Jerome Engel, Eishi Asano, Vwani Roychowdhury, Hiroki Nariai

## Abstract

Epilepsy affects approximately 1% of the global population and is characterized by recurrent seizures arising from large-scale neuronal network synchronisation. However, a unified framework explaining the governing network dynamics both during seizures and in the intervals between them remains elusive. We hypothesise that, analogous to other complex dynamical systems, seizures represent rare, large-scale excursions within a continuum of more frequent, smaller events driven by the same underlying network instability. Using interictal intracranial EEG recordings from 168 patients, we constructed time-resolved, directed high-frequency brain networks and identified two distinct dynamical regimes: a normal (baseline) state and a hypersynchronous state. A small subset of network hubs recurrently drove brief interictal hypersynchronous events, which we term *mini-seizures*. These mini-seizures exhibited scale-free distributions, with overt seizure synchronisation events occupying the extreme tail of the spectrum. In a retrospective cohort analysis treating surgical resection as a network-level perturbation, disruption of these hubs was associated with postoperative seizure freedom. Together, these findings demonstrate that the latent epileptogenic zone—and its clinically relevant postoperative dynamics—can be inferred from recurrent interictal mini-seizure activity alone, supporting an interictal–ictal continuum of epileptic network dynamics.

## Introduction

Epilepsy is one of the most common neurological disorders, affecting approximately 1% of the global population and imposing a substantial health burden worldwide.^1, 2^ Despite major advances in pharmacotherapy, nearly one-third of individuals with epilepsy continue to experience seizures that are resistant to antiseizure medications.^3^ For these patients, focal surgical intervention offers a potentially curative treatment, providing durable seizure freedom and improved long-term survival.^4, 5^ The central target of epilepsy surgery is the epileptogenic zone (EZ), defined as the minimal brain tissue necessary and sufficient for seizure generation.^6^ However, the EZ is a theoretical construct that cannot be directly observed. In clinical practice, it is approximated by the seizure onset zone (SOZ), identified as the brain region showing the earliest sustained electrophysiological change at seizure onset.^6, 7^ Determining the SOZ typically requires days to weeks of intracranial EEG (iEEG) monitoring to capture spontaneous seizures—events that occupy only a minute fraction of the recorded data.^7, 8^ Even when seizures are captured, SOZ localisation may be incomplete or ambiguous, particularly in patients with infrequent seizures or complex propagation patterns.^9^ As a result, more than one-third of patients do not achieve long-term seizure freedom after surgery,^10^ indicating that seizure-based localisation often fails to fully capture the underlying EZ. These limitations highlight a fundamental gap: seizures capture only the extreme tail of epileptic dynamics, while most of the recorded brain activity occurs between seizures and remains underutilised. Despite decades of investigation, how epileptic network dynamics emerge and persist outside overt seizures remains poorly understood. Addressing this gap is critical, as interictal activity constitutes the dominant neurophysiological state of the epileptic brain and may contain latent information about the networks that generate seizures.

A growing body of work has reframed epilepsy as a disorder of distributed brain networks rather than isolated pathological regions.^11, 12^ Network analyses across invasive and non-invasive modalities have demonstrated that epileptic activity involves coordinated interactions among multiple brain regions, with certain nodes exerting disproportionate influence over network behaviour.^13-16^ These findings have motivated the development of network-based frameworks for EZ localisation and outcome prediction. However, most existing approaches characterise baseline or slowly varying network states and are not designed to capture rapid, transient, state-dependent dynamics that may arise during interictal periods. In many complex dynamical systems, pathological behaviour does not emerge as discrete events but instead unfolds across a continuum of activity spanning multiple spatial and temporal scales. These systems frequently exhibit burst-like dynamics, in which small, recurrent fluctuations are punctuated by rare, large-scale excursions driven by the same underlying instability.^17-21^ In such systems, key structural properties can often be inferred from the statistics of small events alone, without direct observation of catastrophic failures.^18^ Neuronal networks are known to exhibit similar scale-invariant dynamics, in which activity fluctuates around an underlying instability threshold and pathological transitions reflect crossings of a critical boundary rather than qualitatively distinct states.^22-26^

We hypothesise that epileptic networks operate under analogous principles, such that brief, recurrent hypersynchronous events during interictal periods reflect transient breakdowns of inhibitory control within the EZ. These events represent scaled-down manifestations of the same network dynamics that culminate in clinical seizures, forming a continuous spectrum from subclinical to overt synchronisation. Here, we test this hypothesis using intracranial EEG recordings from 168 patients across two large epilepsy centres. We develop High-Frequency Synchronisation Network Dynamics (HiSyncDx), a non-linear graph-theoretic framework based on directed high-frequency synchronisation that constructs time-resolved brain networks at second-by-second resolution, building on the inferring connections of networks methodology.^27^ Using this approach, we identify two distinct network states—a normal (baseline) network state and a hypersynchronous network (HSN) state—each exhibiting scale-free features indicative of complex system dynamics. These events are found to be driven by a small subset of hub regions that exert disproportionate influence over network synchronisation. Interictal HSNs occur frequently between overt seizures, span a graded range of magnitudes, and share system-level network architecture with the stronger HSNs observed during seizures, yet remain subclinical; we therefore term them mini-seizures. Much like the frequent microearthquakes that geologists use to map fault zones^18^—events that are typically unfelt by the population—mini-seizures may represent scale-free fluctuations that reveal the latent structure of the epileptic network. Using an unsupervised approach, we identified driver nodes from brief interictal recordings, revealing HSN architectures that replicate ictal dynamics and enabling assignment of an EZ centrality score to each channel—a continuous, data-driven value quantifying its influence within HSNs. Finally, by integrating EZ centrality scores with network flow features, we show that surgical disruption of predicted driver nodes is associated with postoperative seizure freedom and demonstrates high discriminative performance, significantly outperforming models based on seizure onset zone resection alone or spike-associated high-frequency oscillations (mean F1 score, 90% versus 78% and 79%, respectively; P < 0.001). Together, these findings support mini-seizures as a network-level epileptic phenomenon expressed outside overt seizures and identify them as an actionable interictal EEG biomarker to enable precision epilepsy surgery.

## Results

### Emergence of interictal hypersynchronous network states: multi-scale dynamics

We analysed iEEG recordings via subdural grid, strip, or stereotactic EEG (SEEG) from 168 patients with focal drug-resistant epilepsy who underwent invasive monitoring at two tertiary epilepsy centres, followed by surgical resection and postoperative outcome assessment. Full cohort characteristics and inclusion criteria are provided in the Methods (and in Extended Data Table 1). Using our synchronisation-network framework (HiSyncDx), we modelled iEEG as time-resolved, directed high-frequency synchronisation networks to characterise interictal network dynamics.

Interictal EEG is traditionally regarded as a normal state between seizures. Although epileptiform discharges such as spikes and HFOs^28-31^ may emerge during this period, these events are typically analysed as isolated, channel-level phenomena and have shown limited specificity for delineating the EZ.^6, 32-34^ If, however, epilepsy is fundamentally a network disorder, then, pathological synchronisation should recur outside overt seizures as transient, system-level events engaging coordinated interactions across brain regions rather than isolated channels.^11, 12, 14^

To test this hypothesis, we modelled iEEG as time-resolved, directed synchronisation networks constructed from high-frequency activity. This approach is based on the inferring connections of networks (ICON) framework,^27^ which is designed to recover dynamic, directed interactions in large-scale, non-linear systems without imposing stationarity or linearity assumptions. By applying ICON to short, non-overlapping interictal iEEG segments on a second-by-second basis, we generated a sequence of directed graphs that capture rapid changes in synchronisation structure across the recording, consistent with prior observations that epileptic networks undergo fast, state-dependent reconfiguration.^13, 15^

For each temporal window, EEG signals were decomposed into high gamma (50–80 Hz), ripple (80–250 Hz) and fast ripple (250–400 Hz) frequency bands, and pairwise directed synchronisation was estimated between all sampled regions to construct weighted, directed networks (Fig. 1a). Each network snapshot therefore represents the instantaneous large-scale organisation of high-frequency neuronal interactions during interictal activity (see Extended Data Figs. 1 and 2 for justification of the frequency band selection).

**Fig. 1.**
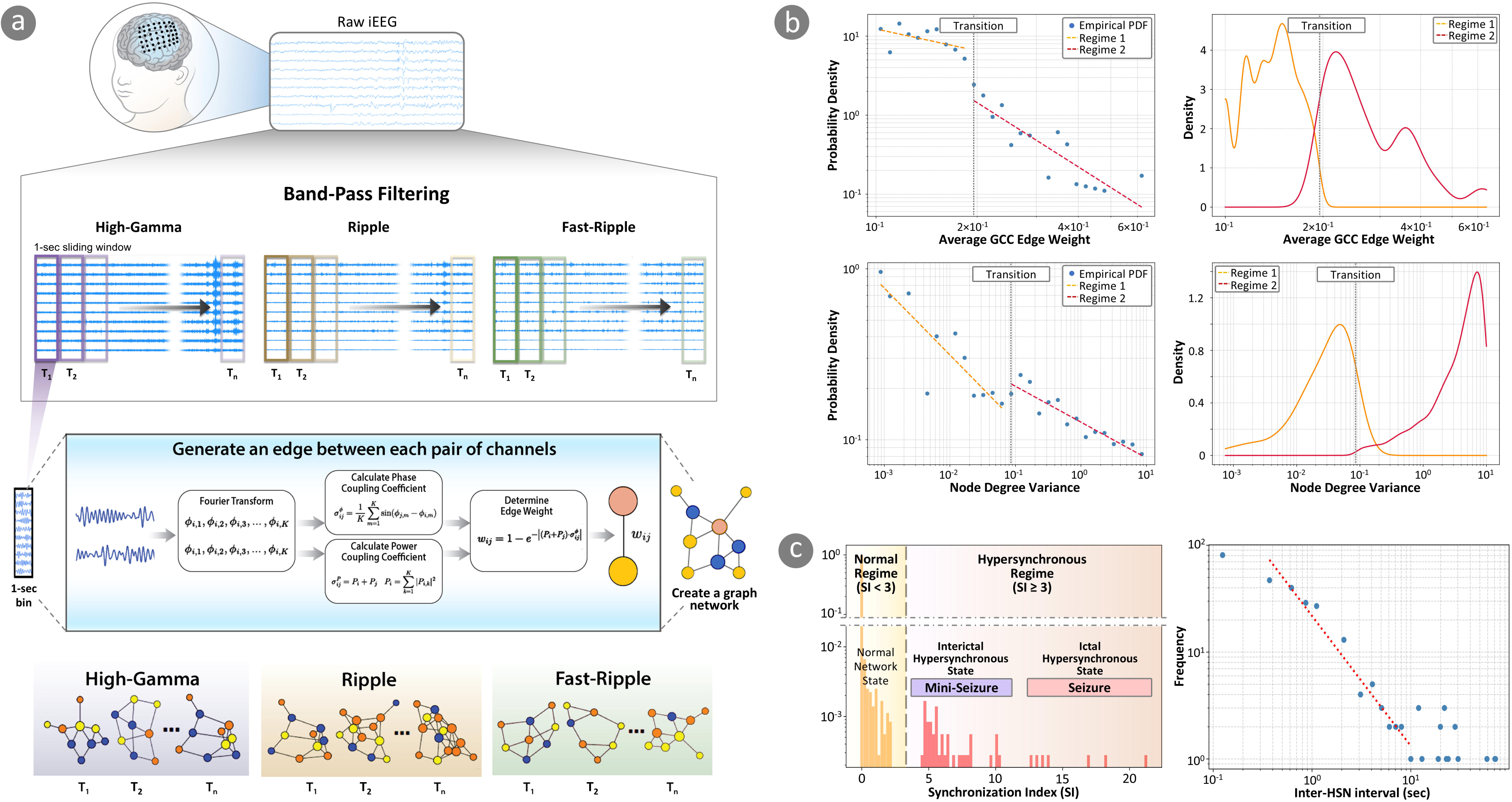
Emergence of two distinct network states in synchronization networks. (a) Schematic overview of synchronization network construction from iEEG. Signals are band-pass filtered into high-gamma (50–80 Hz), ripple (80–250 Hz), and fast ripple (250–400 Hz) bands. For each non-overlapping 1-s window, pairwise power-phase coupling between channels is estimated in the frequency domain to construct a weighted, directed graph, yielding time-resolved synchronization networks for each frequency band. (b) Analyzing the synchronization networks constructed from interictal and ictal iEEG, across the entire cohort, reveals two distinct dynamical regimes in the network metrics space. Distributions of average giant connected component (GCC) edge weight and node degree variance exhibit scale-invariant behavior in both regimes, with a clear transition between the regimes demonstrated by a change in the linear-fit exponent on log–log plots, consistent with bursty dynamics in complex systems. (c) Synchronization index (SI), defined as the product of network synchronizability (Fiedler eigenvalue), average GCC edge weight, and node degree variance, provides a quantitative metric for mining the two network states. The SI distribution is bimodal at the global level, separating a low SI regime corresponding to the normal network state from a high-SI regime corresponding to the HSN state. Notably, the HSN regime itself is bimodal: moderately elevated SI values (3-10) characterize interictal mini-seizures (86890 HSN events), whereas extremely high SI values (> 13) mark ictal HSN (19 overt seizures from three patients, 605 HSN events). Together, these regimes suggest a graded escalation of network organization from baseline to interictal hypersynchrony and, ultimately, ictal hypersynchrony. Moreover, the inter-HSN intervals follow power-law distribution.

**Fig. 2.**
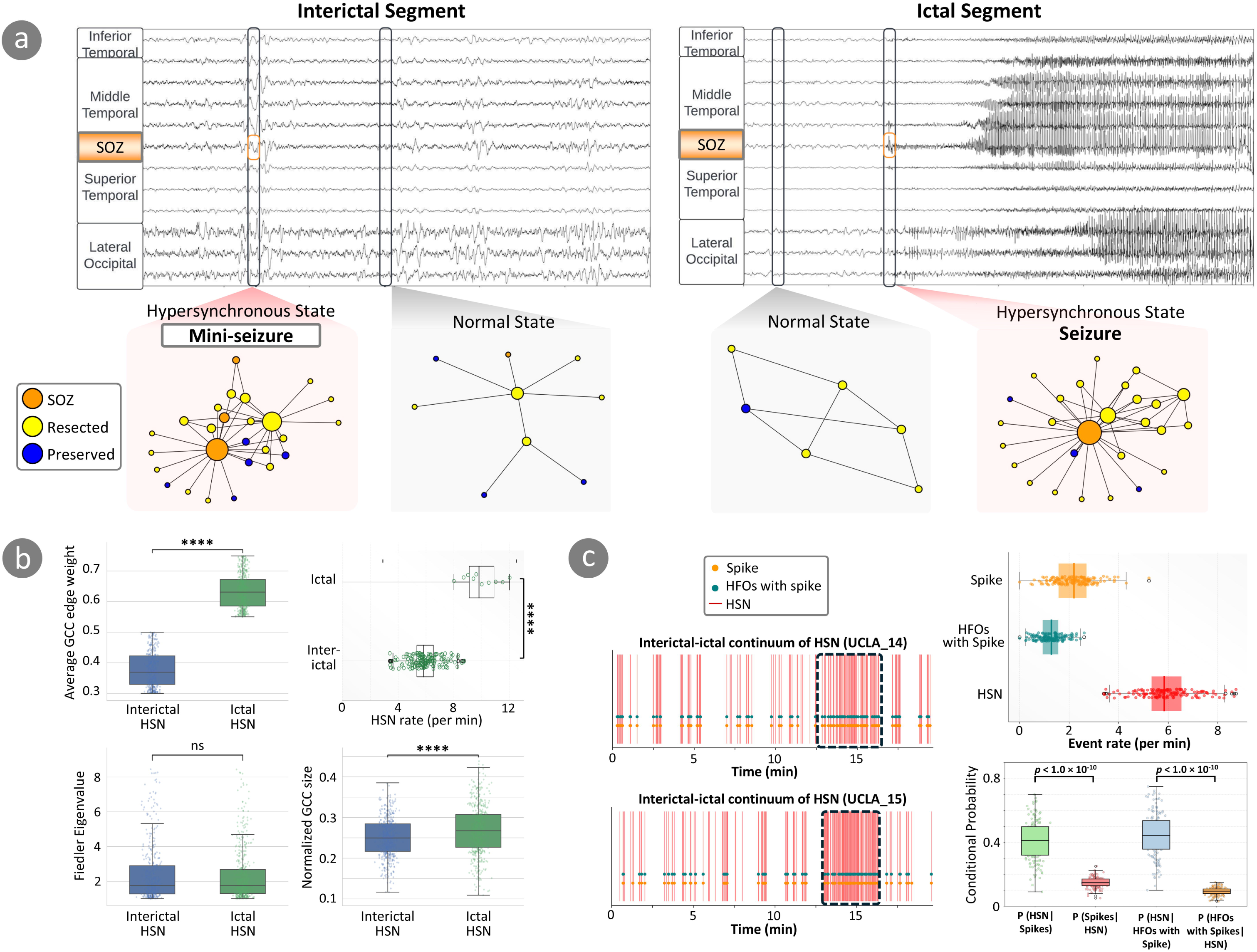
Mini-seizures occur as part of a continuum encompassing similar neurophysiological mechanisms in both interictal and ictal states. (a) Representative iEEG recordings and corresponding synchronization networks from interictal and ictal periods. During interictal segments, mini-seizures emerge transiently with a hub-dominated connectivity pattern involving the SOZ. In contrast, normal interictal states show weak, distributed connectivity. During ictal segments, mini-seizures emerge more frequently with network topology similar to that of interictal mini-seizures but with a greater spatial extent and synchronization strength. Node color denotes SOZ (orange), resected (yellow), and preserved (blue) electrodes. (b) Quantitative comparison between interictal mini-seizures (6061 HSN events) and overt seizures (19 overt seizures from three patients; 605 HSN events). Ictal HSNs exhibit significantly larger GCC size with higher average edge weight compared to interictal mini-seizures, whereas network synchronizability (Fiedler eigenvalue) shows no significant difference. Ictal HSNs occur much more frequently (median 10/min) than interictal mini-seizures (median 5.8/min). (c) Interictal mini-seizures and channel-level discharges may reflect a shared underlying epileptic network process, with mini-seizures providing a more robust readout. Left panels show the timing of mini-seizures, spikes, and spike-associated HFOs in two representative patients across extended recordings. The frequency of HSNs increases during seizures (dashed boxes). Top-right panel shows event rates for spikes, HFOs with spike, and mini-seizures, demonstrating that interictal mini-seizures occur at substantially higher rates than conventional epileptiform activities. Furthermore, conditional probability analysis (bottom-right panel) demonstrates a potentially deeper dynamical dependence between channel-level biomarkers and mini-seizures: whenever a channel-level event occurs (HFOs with spike or spikes) they are predominantly embedded in an mini-seizure whereas a significant number of mini-seizures occur without the presence of any epileptiform activities.

To identify network states characterised by large-scale synchronisation, we quantified global graph properties reflecting complementary aspects of network organisation, including network synchronisability (Fiedler eigenvalue), overall synchronisation strength (average edge weight within the giant connected component), and hub dominance (node-degree variance). Each metric captures a distinct dimension of coordinated network behaviour, spanning integration, coupling strength and hub formation. These measures were combined into a single synchronisation index, enabling objective, data-driven detection and ranking of HSN states within interictal recordings, analogous to approaches used to characterise emergent dynamics in other complex biological and physical systems (see details in the Methods).

Using this framework, we observed the spontaneous emergence of transient HSN states during interictal periods. Analysis of global network metrics revealed two distinct dynamical regimes—a normal (baseline) regime and a hypersynchronous regime—separated by a transition boundary in synchronization index space. Relative to baseline activity, HSN states were characterised by concurrent increases in network synchronisability, strengthened connectivity within the giant connected component, and pronounced hub formation, indicating coordinated, system-level reorganisation rather than focal channel-specific changes (Fig. 1b). Network metrics within the hypersynchronous regime exhibited scale-free, power-law distributions, consistent with bursty dynamics across multiple temporal and spatial scales. Analysis of the synchronisation index across all interictal recordings revealed a bimodal distribution, separating baseline network states from a distinct hypersynchronous regime (Fig. 1c and Extended Data Fig. 3). Importantly, HSN states occupied an intermediate range of synchronisation values between baseline activity and full ictal seizures, indicating a graded hierarchy of network states rather than a binary interictal–ictal transition.

The hypersynchronous regime further exhibited non-Gaussian, heavy-tailed distributions across multiple network metrics. Synchronisation strength, network synchronisability and degree variance all followed approximate power-law scaling within HSN states, consistent with scale-free dynamics observed in other complex systems (Fig. 1b).^17-20^ In contrast, baseline interictal activity lacked hub dominance and large-scale integration and showed markedly different statistical structure.

Together, these findings show that interictal EEG is not a stationary baseline but is punctuated by brief HSN events intermediate between baseline and ictal states. Their hierarchical, scale-free organisation indicates an intrinsic epileptic network process rather than stochastic fluctuations or isolated discharges. We therefore refer to these events as interictal mini-seizures, which are distinguished from channel-level HFOs or microseizures^23, 24^ by transient, system-level network reorganization with hub dominance.

### Hypersynchronous network states are a continuum bridging interictal and ictal network states

The identification of recurrent interictal HSN states (mini-seizures) raises a fundamental question: how are these events related to the large-scale network dynamics observed during seizures? If interictal mini-seizures reflect intrinsic properties of epileptic networks rather than incidental fluctuations, their organisation should resemble—and potentially anticipate—the network architecture expressed at ictal period.

To address this question, we compared network architecture across interictal and ictal EEG segments using the same synchronisation-network framework. To summarize, during interictal periods, mini-seizures emerged transiently, characterised by brief increases in large-scale network connectivity and hub engagement (Fig. 2a). Strikingly, the network characteristics observed during interictal mini-seizures closely resembled that seen at seizure onset, with overlapping patterns of directed connectivity and hub-like structure despite differences in spatial extent and synchronisation strengths (Fig. 2a/2b).

We next examined the temporal structure and recurrence of mini-seizures. Across the entire cohort, these events were frequent, with a median occurrence rate of 5.8 events per minute, indicating that hypersynchronous states are a common feature of interictal network dynamics rather than rare anomalies. Analysis of the intervals between successive events revealed an approximate power-law distribution spanning nearly two orders of magnitude, a hallmark of self-organised complex systems (Fig. 1c).^17, 26^ This scale-free temporal organisation suggests that hypersynchronous events arise from intrinsic network dynamics capable of generating activity across a broad range of magnitudes and time scales, encompassing overt seizures.

HSN states also occur at higher rates than epileptiform discharges, such as spikes HFOs with spikes (Fig. 2c); moreover, their conditional co-occurrence statistics suggest that HSNs and channel-level discharges may reflect a shared underlying epileptic network process, with HSNs providing a more robust readout.

Together, these findings demonstrate that interictal mini-seizures and seizures are not distinct categories of activity but instead represent different expressions along a continuous spectrum of epileptic network synchronisation. We demonstrated that mini-seizures occupy the low-amplitude, short-duration end of this spectrum, whereas seizures correspond to its rare, high-amplitude, sustained extreme.

### Unsupervised discovery of nodes driving hypersynchronous network states

If interictal mini-seizures reflect intrinsic epileptic dynamics, they should be initiated and sustained by a subset of nodes that repeatedly are the hubs driving network synchronisation. Collectively the set of such driver nodes (we will use hubs and driver nodes interchangeably) would be expected to serve as the network-level substrate of the EZ.

We examined node-level network features during interictal mini-seizures (Fig. 3a) and asked whether they could be used to identify driver nodes in a data-driven, unsupervised manner. For each channel, we quantified multiple graph-theoretic features capturing complementary aspects of directed network influence, including in-degree, out-degree, eigenvector centrality, and metrics scaled by network synchronisability (i.e. Fiedler eigenvalues). These features were summarised over time using higher-order statistics to capture the non-Gaussian, heavy-tailed dynamics observed during hypersynchronous states (see Methods).

**Fig. 3.**
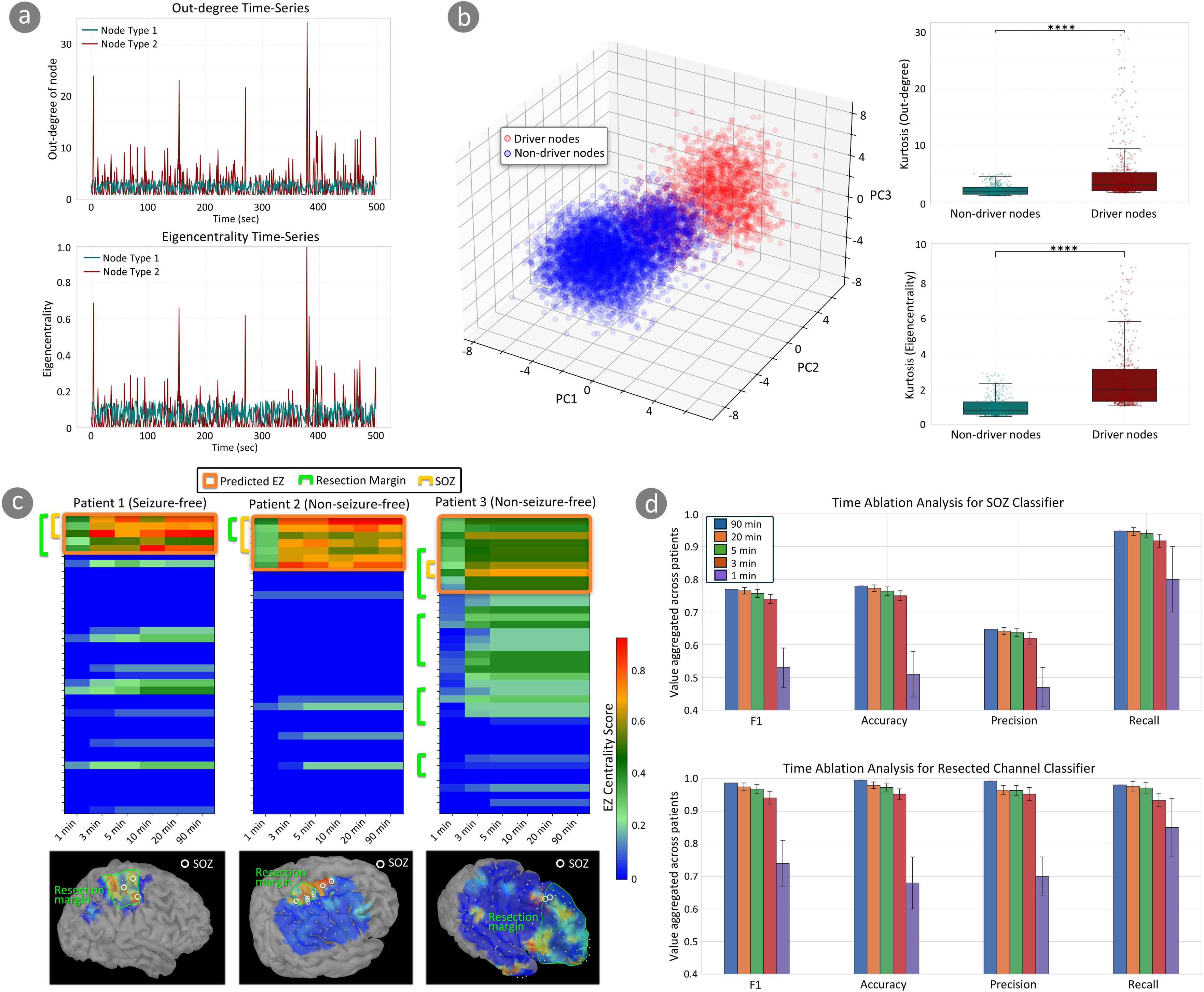
Unsupervised identification of driver nodes of interictal mini-seizures enables high-confidence epileptogenic zone (EZ) localisation. (a) Some of the nodes exhibit persistent and significantly greater directed influence during interictal mini-seizures than other nodes. In the plot, node type 2 has repeated high values of out-degree and eigenvector centrality—thus acting as a driver or hub node—while the other node remains consistently as a peripheral one. This motivates, a selection of a set of features, such as the kurtosis of out-degree (see Methods) etc. that could distinguish between such classes of nodes. (b) Clustering algorithm applied to the node-level feature vectors discovered two classes of nodes in the interictal HSN. The cluster associated with higher mean network synchrony—quantified by average pairwise synchronization within the cluster during HSN events—was designated as the driver cluster. As illustrated in the box plot, the nodes in the driver cluster exhibit significantly greater directed influence during interictal HSNs. For each channel, the EZ centrality score was defined as the inverse of its Euclidean distance to the centroid of this driver cluster, and then normalized to a value in the interval [0,1]. The separation between these clusters is even evident in a 3-dimensional space. Nodes with EZ centrality score ≥ 0.5 are colored in red. (c) Spatial correspondence between predicted driver nodes and surgical outcome in representative patients. Heatmaps of EZ centrality scores for three patients are shown with rows representing the channels and columns representing the duration of the iEEG epoch used to compute the EZ centrality. Patients whose resection margins encompassed all channels with high EZ centrality score (≥ 0.5) achieved seizure freedom (left), whereas patients with preserved high-centrality driver nodes experienced seizure recurrence (middle and right). In the rightmost example, resection included the clinically annotated SOZ, yet additional predicted driver nodes outside the SOZ were preserved, suggesting incomplete disruption of EZ. (d) Robust performance of driver-node–based localisation using short interictal recordings. For an iEEG segment of given duration, we compute EZ centrality scores for all channels and convert them into binary predictions (EZ vs. Non-EZ): any channel with an EZ centrality score ≥ 0.5 is designated as EZ. We then quantify performance by comparing the predicted EZ labels against two ground-truth definitions: (i) the clinically annotated seizure onset zone (SOZ) (ii) the set of resected channels. High performance is achieved with iEEG recordings as short as 3 minutes, with mean F1 scores of approximately 0.77 for SOZ localisation and 0.97 for resected-channel identification, indicating that brief interictal segments contain sufficient information to delineate clinically relevant EZ.

Applying clustering to these node-level feature representations revealed two well-separated clusters across patients and recordings (Fig. 3b). One cluster was characterised by consistently elevated out-degree and eigenvector centrality, indicative of nodes that actively drive synchronisation across interictal period, whereas the other cluster comprised of nodes exhibiting lower influence and more distributed connectivity. We therefore designated these groups as driver nodes and non-driver nodes, respectively. Importantly, this separation emerged without the use of seizure annotations, anatomical priors, or outcome information, indicating that driver nodes can be identified directly from intrinsic interictal network dynamics. Such driver nodes consistently exhibited spatial organisation centred on regions involved in SOZ and presumed EZ (as quantified in Fig. 3(d) and Extended Data Fig. 4).

Significantly, we found that the driver nodes switch roles during normal network states, where its edges get directed inwards (suggesting functional suppression), reversing its outward direction during mini-seizures (Extended Data Fig. 5). This pattern of switching roles is consistent with SOZ behavior described in the literature.^8, 16^ These findings establish a mechanistic link between network hubs identified during interictal mini-seizures and the initiation of pathological synchronisation.

To translate these node-level distinctions into a continuous, interpretable measure of epileptogenicity, we defined an EZ centrality score for each channel, reflecting its proximity to the driver-node cluster in feature space. This score provides a graded estimate of each channel’s likelihood of acting as a network driver and, by extension, belonging to the EZ. Although the full computational definition is described in the Methods, conceptually the EZ centrality score captures how strongly a given channel expresses the multivariate network signature of driver nodes. The decision threshold of 0.5 was selected based on cross-validated optimization (Extended Data Fig. 6).

Mapping EZ centrality scores back onto individual patients revealed a close relationship between predicted driver nodes and clinically relevant regions. In representative cases, seizure-free patients showed near-complete resection of channels with high EZ centrality scores, whereas patients with persistent seizures retained subsets of high-centrality driver nodes despite resection of the clinically annotated SOZ (Fig. 3c). Notably, in some cases, preserved driver nodes lay outside the SOZ, suggesting that conventional SOZ-based approaches may fail to capture the full extent of EZ.

At the group level, driver nodes identified from as little as three minutes of interictal EEG showed strong spatial overlap with both physician-annotated SOZs and resected regions (Fig. 3d). Across patients, EZ centrality–based predictions achieved a mean F1 score of approximately 0.77 for SOZ localisation and 0.97 for resected-channel identification, demonstrating that brief interictal recordings contain sufficient information to delineate clinically meaningful EZ.

### Disruption of network drivers predicts postoperative seizure freedom

If driver nodes identified from interictal network dynamics represent the causal substrate of seizures, then surgical disruption of these nodes—and the network interactions they sustain—should predict postoperative outcome. Surgical resection therefore provides a natural perturbation experiment to test the functional relevance of EZ centrality and network drivers.

We introduced the EZ centrality resection ratio (Fig. 4a and Methods), a scalar metric quantifying the proportion of predicted EZ removed at surgery. This measure is analogous to the HFO resection ratio used in seizure outcome prediction based on channel-level biomarkers. However, a single scalar ratio does not capture how resection reshapes network interactions. We therefore developed a network flow framework to quantify directed high-frequency synchronisation between predicted EZ and preserved regions (Fig. 4a). Channels were partitioned into resected EZ nodes, preserved EZ nodes, and preserved non-EZ nodes according to EZ centrality scores and surgical margins. Synchronisation flow was then computed as the time-averaged sum of directed edge weights between node sets, yielding a continuous and interpretable measure of residual coupling between preserved EZ nodes and the surrounding network (see Methods).

**Fig. 4.**
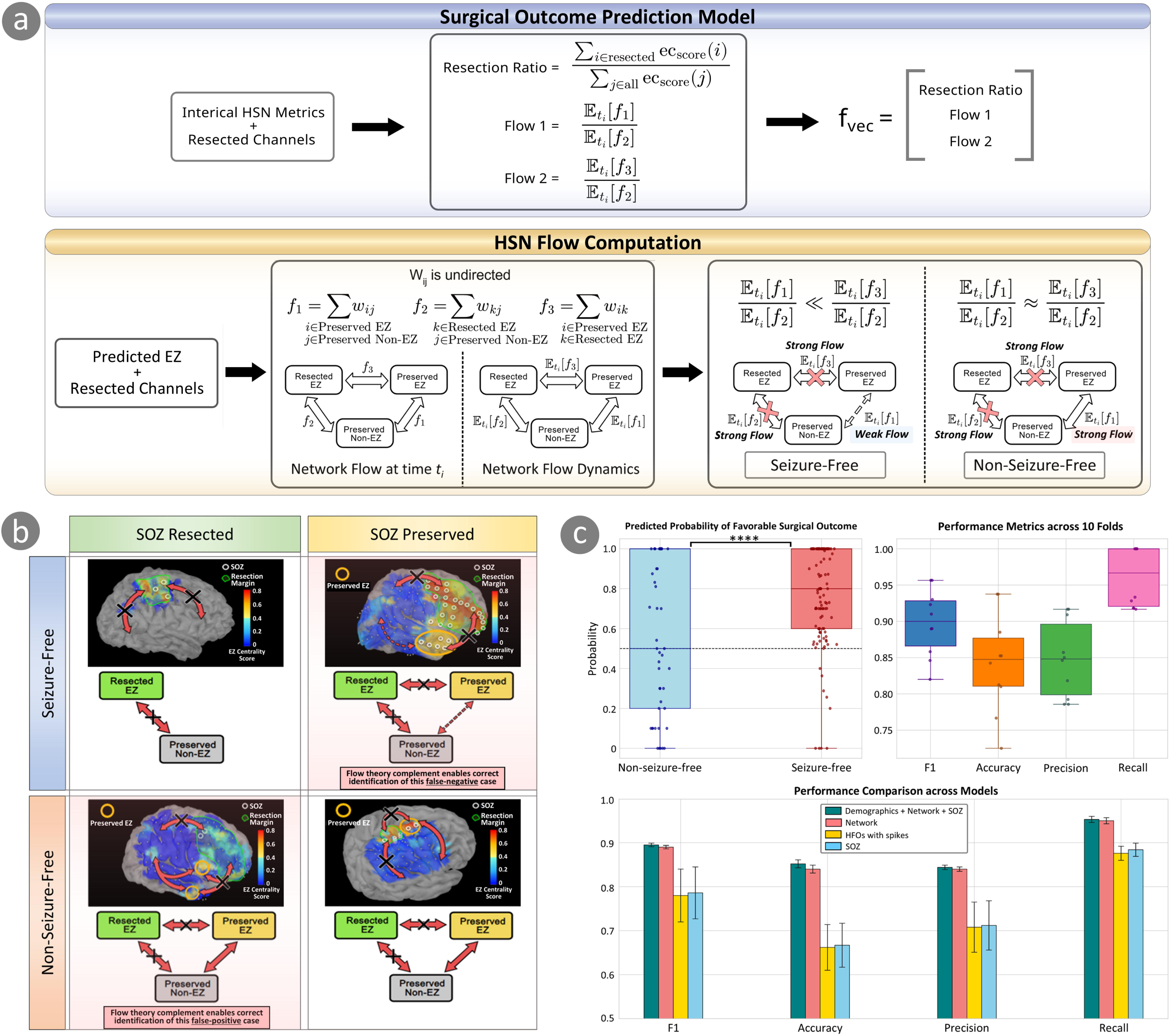
Network-based model accurately predicts postoperative seizure outcome. (a) Schematic of the surgical outcome prediction model incorporating interictal mini-seizure metrics. For each patient, EZ centrality resection ratio is used as the first feature. For flow computations, electrodes are partitioned based on EZ centrality scores and surgical resection status into three non-overlapping sets: resected EZ, preserved EZ, and preserved non-EZ. Undirected synchronization flow between these regions is quantified over time to capture how driver nodes remain coupled to, or isolated from, the preserved brain network. Conceptual flow patterns associated with seizure-free and non–seizure-free outcomes are illustrated. (b) Representative patient examples illustrating how incorporating network flow resolves apparent discrepancies between EZ localization and surgical outcome—discrepancies that arise when EZ-outcome alignment is assessed solely using resection ratio. As expected, complete resection of high EZ-centrality nodes is associated with seizure freedom (top left). Similarly, preservation of EZ nodes that remain strongly coupled to non-EZ regions predicts seizure recurrence (bottom right). However, patients can still achieve seizure freedom despite preservation of the clinically annotated seizure onset zone (SOZ)—also predicted as being part of EZ by HiSyncDx—if they are only weakly coupled to the remaining network (top right). Conversely, complete SOZ and a large fraction of predicted EZ resection does not guarantee seizure freedom if nodes with strong residual network coupling are preserved (bottom left). (c) Cross-validated performance of surgical outcome prediction models. (i) Predicted probability of seizure freedom for individual patients using the network-based model incorporating flow; each point represents one patient, with surgical outcome indicated by color. The dotted line denotes the classification threshold. (ii) Distributions of performance metrics across 10-fold demonstrate robust generalization, with a high mean F1 score (89.8%) and low variance (0.002) across folds. (iii) Comparative evaluation across four models shows that the synchronization network–based model incorporating flow achieves superior performance (mean F1 = 89.8%) relative to models based on HFOs with spike (mean F1 = 77.6%) or SOZ resection status alone, the current clinical standard (mean F1 = 78.0%), with consistent gains across F1 score, accuracy, precision, and recall.

This framework yields clinically interpretable scenarios. Complete resection of high-centrality EZ nodes disrupts outgoing synchronisation flow, effectively isolating EZ from the remaining network. In contrast, preservation of EZ driver nodes—particularly when they retain strong connectivity to non-EZ regions—maintains the capacity for large-scale synchronisation and seizure generation. Representative patient examples illustrate how incorporating network flow resolves apparent discrepancies between resection extent and outcome, correctly identifying false-positive and false-negative predictions based on the EZ centrality resection ratio alone (Fig. 4b).

A random forest classifier was chosen for its ability to capture non-linear interactions among network features without strong parametric assumptions. Predictor variables included the EZ resection ratio, measures of residual synchronisation flow between preserved EZ and non-EZ regions, and flow between resected and preserved EZ regions, providing a quantitative representation of how surgical intervention alters network connectivity. The model was trained and evaluated using stratified cross-validation across the full cohort of 163 patients, with seizure freedom at one year as the primary outcome (see Methods).

The network-based model demonstrated robust predictive performance, achieving a mean F1 score of 0.90, with high recall and low variance across cross-validation folds (Fig. 4c). This performance significantly exceeded that of models based on SOZ resection alone (mean F1 ≈ 0.78) or spike-associated HFOs (mean F1 ≈ 0.79; P < 0.001). Importantly, incorporating network flow features improved discrimination in cases where EZ tissue was partially preserved but functionally disconnected, highlighting the added value of modelling network interactions rather than relying solely on anatomical resection.

Together, these findings demonstrate that by integrating unsupervised driver-node identification with network flow modelling, this framework provides a mechanistically grounded, clinically actionable approach to predicting surgical outcomes from brief interictal recordings.

## Discussion

### A continuous, cross-scale spectrum of epileptic network synchronization

Our results support a view of epilepsy as a dynamic, multiscale network disorder in which pathological synchronisation emerges continuously rather than episodically.^11, 12^ The observation that interictal mini-seizures follow power-law temporal statistics and scale smoothly into ictal events places epilepsy within a broader class of bursty complex systems that generate frequent small events and rare large ones under shared governing dynamics.^19-21, 26^ In such systems, global structure and stress distributions can be inferred from the statistics of small events alone, without direct observation of catastrophic failures—a principle widely exploited in seismology to map fault zones from microearthquakes.^18^

Analogously, we show that epileptic networks continually generate transient hypersynchronous events that reflect latent network instability, like neural fragility.^35^ These mini-seizures are not random fluctuations, but structured network states characterised by hub-dominated topology, increased synchronisability, and scale-free organisation. The graded increase in event frequency and persistence leading up to seizures further supports the existence of an interictal–ictal continuum of network synchronisation, consistent with prior observations of progressive network recruitment preceding seizure onset.^23, 24, 35^ This continuum is also consistent with large-scale EEG studies demonstrating graded transitions between interictal, ictal, and peri-ictal rhythmic and periodic patterns during expert EEG interpretation, rather than sharp categorical boundaries between states.^36^ This framework provides a unifying, system-level description of epileptic activity that bridges traditionally distinct interictal and ictal states.

Experimental and theoretical studies have suggested that neuronal networks operate near criticality, balancing excitation and inhibition to maximise dynamic range and responsiveness.^22, 25^ Our findings extend this concept to human epileptic networks, demonstrating that pathological synchronisation emerges as a quantitative deviation along a continuous dynamical spectrum rather than as a qualitatively distinct state. From this perspective, seizures represent extreme excursions of an otherwise persistent network process.

### Relation to existing network-based frameworks

Several network-based frameworks have advanced understanding of epileptic networks. Neural fragility characterises the susceptibility of brain networks to destabilisation and has demonstrated utility for identifying the SOZ, but relies on linear system approximations and typically requires ictal data for model training.^35^ Source–sink models and the interictal suppression hypothesis emphasise baseline inhibitory restraint of EZ during interictal periods, supported by network-level evidence across modalities.^14, 16, 37^

HiSyncDx complements and extends these approaches in several important ways. First, it directly models non-linear, non-stationary network dynamics without imposing linearity or extended temporal averaging, enabling detection of rapid, transient synchronisation events that may be overlooked by static or baseline-focused analyses. Second, rather than characterising the EZ solely in a suppressed or baseline state, HiSyncDx identifies network hubs in their active, source-like configuration during hypersynchronous events. Third, the framework can operate entirely on interictal data, obviating the need to capture seizures for network inference. Consistent with this perspective, recent work has shown that machine-learning analysis of interictal iEEG alone can delineate EZ and predict postoperative seizure outcomes without requiring ictal data, further supporting the view that clinically actionable information about epileptic networks is embedded in interictal dynamics.^38^

Importantly, these approaches need not be viewed as competing. Instead, they may reflect complementary projections of the same underlying network architecture across distinct dynamical regimes. As illustrated Extended Data Fig. 5, the driver nodes in our framework switch roles between being a sink and a source: in normal synchronization state its functionality is suppressed and the edges are pointed inwards; however, in HSN states the suppression mechanism fails and it drives synchronisation with its edges pointed outwards. Baseline suppression, pre-ictal instability, transient interictal hypersynchrony, and sustained ictal synchronisation may represent different expressions of a unified epileptic network process. HiSyncDx specifically targets the transient regime in which pathological synchronisation becomes briefly expressed but does not yet generalise into a full seizure.

### Clinical and translational implications

From a translational standpoint, interictal mini-seizures represent a network-level interictal EEG biomarker with graded dynamical properties rather than a binary event marker. Unlike conventional channel-based markers such as spikes or HFOs, mini-seizures are defined at the network level, capturing coordinated interactions across regions rather than isolated local events. This distinction is critical, as channel-based HFOs have shown limited specificity and inconsistent predictive value for surgical outcome, in part because they can arise in physiologically normal cortex and do not explicitly encode network organisation.^32-34^ Some approaches leverage spike-based or HFO-based connectivity to localise EZ; however, these methods typically depend on prior detection of channel-level epileptiform events (spikes or HFOs) before network construction.^39, 40^

By contrast, mini-seizures encode both where pathological synchronisation emerges, centred on driver nodes, and how the HSN is organised. The unsupervised identification of driver nodes provides a principled, data-driven representation of the EZ that extends beyond the SOZ. Notably, we show that these drivers can be detected from minutes of interictal iEEG and that their surgical disruption predicts seizure freedom more accurately than SOZ resection or HFO-based models.

The integration of network flow enhances clinical interpretability by explaining why some patients achieve seizure freedom despite incomplete resection of predicted EZ tissue—namely, because residual driver nodes become functionally disconnected from the preserved network. This network-centric perspective reframes epilepsy surgery as an intervention on pathological network interactions rather than solely on anatomical regions.

### Limitations and future directions

Although our findings identify stable network drivers whose disruption predicts outcome, causality is inferred indirectly through surgical perturbation rather than direct experimental manipulation. Several additional limitations warrant consideration. First, our analyses relied on macroelectrode iEEG recordings, which cannot resolve single-neuron or microcircuit-level dynamics. Integrating microelectrode and macroelectrode recordings may help link network-level mini-seizures to cellular mechanisms of excitation–inhibition imbalance.^41-43^ Second, we focused on short interictal segments acquired early during monitoring. While this demonstrates feasibility for rapid EZ localisation, longer recordings may reveal additional structure in mini-seizure dynamics, including circadian variation and modulation across sleep–wake and vigilance states.^44, 45^

Future studies should evaluate whether interictal mini-seizure–guided network mapping can inform resection planning and whether targeted disruption of predicted network drivers improves outcomes relative to current standard-of-care approaches. Beyond EZ localisation, this spatio-temporal network framework may also enable a broader redefinition of surgical planning. Once prospectively validated, it could support virtual resection strategies^46, 47^ in which dynamical network models are used to simulate the impact of candidate surgical interventions, estimating not only the probability of seizure freedom but also potential effects on cognitive and functional networks. Realising this vision will require integration of interictal network dynamics with stimulation-based mapping and eloquent cortex localisation routinely performed during invasive monitoring, thereby extending the framework from EZ identification to patient-specific modelling of both pathological and functional brain networks.

## Conclusion

In summary, this work introduces interictal mini-seizures as a window into the latent dynamics of epileptic networks. By framing epilepsy as a continuously active, multiscale network process and by leveraging frequent interictal events rather than rare seizures, HiSyncDx provides a unified neuroscience and translational framework for understanding, localising, and ultimately intervening in pathological brain synchronisation.

## Materials and methods

### Patient cohort and dataset

We conducted a multi-institutional retrospective cohort study including pediatric-onset (≤ 21 years) patients with focal drug-resistant epilepsy (DRE) who underwent invasive iEEG monitoring and subsequent resective surgery. Patients were recruited from UCLA Mattel Children’s Hospital (August 2016–December 2023) and Children’s Hospital of Michigan, Detroit (January 2007–May 2018). Inclusion criteria required: (1) video-iEEG recording with subdural grid, strip electrodes, or SEEG; (2) a sampling rate of at least 1,000 Hz; (3) at least one artifact-free 5-minute slow-wave sleep epoch recorded at least two hours apart from seizures; (4) completion of resective surgery; and (5) documented postoperative seizure outcomes at one year or more. Patients were excluded if they underwent epilepsy surgery without iEEG, had massive brain malformations or previous surgeries preventing accurate anatomical identification, or lacked postoperative outcome data. The study protocol was approved by the institutional review boards at UCLA and Wayne State University, and written informed consent was obtained from patients or guardians.

A standardized presurgical evaluation was performed for all subjects, including inpatient video-EEG monitoring, high-resolution (3.0 T) MRI, and 18-fluoro-deoxyglucose positron emission tomography (FDG-PET) with MRI–PET co-registration. Clinicians identified SOZs as regions exhibiting the earliest sustained rhythmic activity at seizure onset.^6^ For the ictal data, six patients who provided seizures during the first night were analyzed. The extent of resection was guided primarily by the SOZ while avoiding unacceptable neurological deficits. Postoperative outcomes were classified according to ILAE criteria, with class I indicating seizure-free status and all other classes indicating seizure recurrence at ≥ 12 months.

iEEG recordings were obtained using Nihon Kohden systems (Irvine, CA, USA) at 1,000 Hz in Detroit and 2,000 Hz in UCLA. Platinum grid electrodes (10 mm intercontact distance) and depth/SEEG electrodes (platinum, 5 mm intercontact distance), were surgically implanted. Electrode contacts were co-registered to patient-specific three-dimensional (3D) brain surfaces derived from preoperative high-resolution MPRAGE T1-weighted images. FreeSurfer software was used to generate cortical surfaces, with manual correction where pial surfaces were inaccurately detected. For spatial normalisation and cross-institutional harmonisation, electrodes from Detroit were converted to MNI space and combined with UCLA data using Brainstorm software.^48^ Electrodes outside the brain or containing significant artefacts were excluded from further analysis. The resection status of each electrode (resected vs preserved) was confirmed using post-resection MRI and intraoperative photographs.^48^

For preprocessing, all EEG signals were referenced using a bipolar montage. UCLA data were resampled to 1,000 Hz to match the Detroit recordings. A 2-Hz stopband band-reject filter was applied to remove 60-Hz line noise and its harmonics. Artifact-free interictal iEEG epochs of 5–90 minutes were extracted from the first night of monitoring, prior to medication tapering, for subsequent analysis. Notably, the majority of the Detroit iEEG recordings were 5 minutes in duration, whereas the majority of the UCLA recordings were 90 minutes (details in Extended Data Table 1). Ictal iEEG data were analysed from a subset of the UCLA cohort (six patients), comprising all available artifact-free seizures with clearly defined electrographic onset during the study period (details in Extended Data Table 2). These ictal recordings were used to characterise ictal network architecture and compare it with interictal mini-seizures; they were not used for model training or outcome prediction. For traditional channel-based EEG marker detection (interictal spikes and spike-associated HFOs), we used our previously validated software package, pyHFO.^49, 50^

### HiSyncDx: constructing high-frequency synchronization network (Fig 1a)

#### Network construction overview

For a given frequency band, we construct a directed, weighted, time-varying synchronization network from iEEG following the inferring connections of networks (ICON) framework.^27^ First, the iEEG is partitioned into non-overlapping 1-second segments. For each segment, we build a directed weighted graph with N nodes, where N equals the number of iEEG channels. Pairwise edge weights are derived from Fourier-based power- and phase-coupling measures, and edge directionality is determined from the phase-coupling term. Finally, to retain only the most salient interactions, we apply a dynamic threshold that preserves edge weights in the top 10th percentile (Extended Data Fig. 3).

#### Directed edge weight construction

For each channel, we compute its power and phase spectrum using Fast Fourier Transform (FFT). After obtaining its power spectrum, we isolate the given frequency band and set the magnitude of each frequency component outside the given band to zero.

Let *Pi* be the sum of the squared power spectrum of the *i^th^* node and is given by

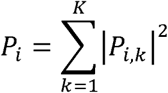

where *K* is the number of discrete frequencies in the given frequency band. Then, power-related coupling coefficient between nodes *i* and *j* is given by

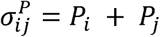

Phase-related coupling coefficient between node *i* and *j* is given by

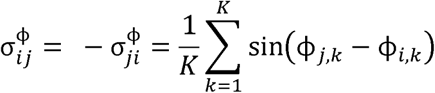

where *K* is the number of discrete frequencies in the given frequency band.

The synchronous coefficient between node *i* and *j*, *d_ij_* is given by

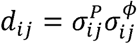

From the above equation, it can be observed that the synchronous coefficient is the product of the power and phase-related coefficient. Then, we compute the edge weight between node *i* and *j*, *w_ij_*, as

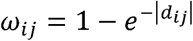

The total phase in node *j* is given by

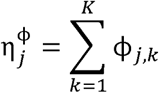

where *K* is the number of discrete frequencies in the given frequency band. Analogously, the total phase in node *i* is given by

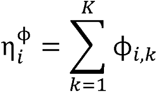

If there is a greater total phase in node *j* than in node *i*, that is

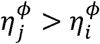

then the weighted edge points from node *i* to node *j* (*i* → *j*). If there is a greater total phase in node *i* than node *j*, that is

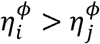

then the weighted edge points from node *j* to node *i* (*j* → *i*).

### Computing network synchronization index, node properties, and other characteristics of high-frequency synchronisation networks

For each of the three frequency bands high gamma (50–80 Hz), ripple (80–250 Hz), and fast ripple (250–400 Hz), HiSyncDx outputs a sequence of T networks (assuming one second-long segments) where T is the duration of the iEEG recordings. To explore the structure and the dynamics of the networks, we use a mix of global (network level) and local (node level) graph theoretic properties. To capture such events in the network level dynamics, we use measures such as (i) Fiedler eigenvalue: higher Fiedler eigenvalue implies higher synchronizability of the networks, (ii) edge weights: larger edge weight implies stronger synchronization strength and (iii) Node degree variance: higher variance implies the presence of hubs. We propose a data-driven computational metric to quantitatively identify synchronization networks that have high values for each of the metrics, referred to as Hypersynchronous Network State (HSN). We refer to this metric as the Network Synchronization Index (SI), which is the product of the three global measures stated above. The metric not only assists in identifying HSNs but also provides a potential framework for ranking the network states and unearthing interictal time stamps of interest in an unsupervised way. In addition to identifying HSNs, we also identify the drivers of HSNs using a set of node-level features like in-degree, out-degree, scaled in and out degrees by Fiedler eigenvalue and eigencentrality scores.

### Directional co-occurrence between HSN and channel level events (Fig 2c)

For each patient (n = 168), we quantified directional co-occurrence between HSNs (mini-seizures) and conventional interictal events (spikes and spike-associated HFOs). HSN occurrences were defined as 1-second windows identified as the HSN state based on the synchronization index. Spikes and spike-associated HFOs were detected at the channel level and aligned to this common windowing. For each window, we defined a binary “event present” indicator for spikes (or spk-HFOs) as 1 if **any** channel exhibited at least one detected spike (or spk-HFO) within that window, and 0 otherwise. Using these window-level binary sequences, we computed patient-specific conditional probabilities in both directions: P(*HSN|spike*) as the fraction of spike-present windows labeled as HSN, and P(spike∣HSN) as the fraction of HSN windows that were spike-present; the same procedure was repeated for spk-HFOs to obtain P(HSN∣spk-HFO) and P(spk-HFO∣HSN). Patient-level conditional probabilities were summarized across the cohort using boxplots, and differences between directions were assessed across patients using a two-sided paired statistical test (p values shown in Fig 2c).

### Unsupervised EZ discovery: defining EZ centrality score

A purely unsupervised framework was developed to estimate an EZ centrality score for each electrode channel, reflecting its relative influence on HSN dynamics. At each time point, five graph-theoretic measures—in-degree, out-degree, eigenvector centrality, and in-and out-degree scaled by the Fiedler eigenvalue (a measure of network synchronisability)—were computed to capture complementary aspects of directed connectivity and network integration.

Each feature was summarized over time using four statistical moments (mean, variance, skewness, and kurtosis), yielding a 20-dimensional feature vector per channel per frequency band (see Extended Data Figure 7 for the ablation analysis of each feature). Feature vectors across the three frequency bands were concatenated to form a 60-dimensional embedding for each channel.

To identify dominant network patterns, K-Means clustering with k=2 was applied to the channel embeddings, partitioning channels into two groups. The cluster exhibiting higher mean network synchrony---quantified as the average pairwise synchronization among channels within the cluster during HSN events---was designated as the driver cluster. For each channel i, we computed its Euclidean distance d_i_ to the driver-cluster centroid and converted this distance into a normalized inverse-distance--based EZ centrality score bounded in [0,1] as

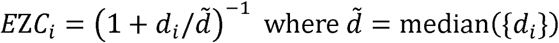

is the median distance across all channels in that recording. Under this mapping, channels closer to the driver centroid receive scores approaching 1, while channels farther away approach 0. This formulation provides a continuous, data-driven measure of each channel’s proximity to the putative EZ, enabling unsupervised identification of network drivers directly from intrinsic topological features without reliance on labeled data or prior anatomical constraints.

### Network flow dynamics

We partitioned the interictal network into three non-overlapping regions based on EZ centrality scores and surgical resection information: resected predicted EZ channels (EZ centrality score > 0.5 that were resected), preserved predicted EZ channels (EZ centrality score > 0.5 that were not resected), and preserved non-EZ channels (EZ centrality score < 0.5 that were not resected). The decision threshold of 0.5 was selected based on cross-validated optimization (Extended Data Fig. 6).

To quantify directed network interactions between these regions, we computed flow from the high-frequency synchronisation networks derived using HiSyncDx. Specifically, flow between node sets was defined as the sum of directed edge weights ω== connecting nodes across regions:

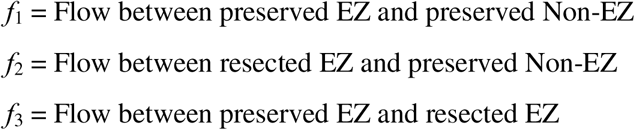

with

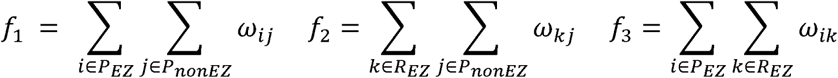

where P_EZ_ = Preserved EZ, P_nonEZ_ = Preserved non-EZ, R_EZ_ = Resected EZ

Note that such a partitioning might not always exist, when there is no preserved EZ, in such cases we set *f*_1_ = *f*_2_ = *f*_3_ = 0. To summarize network flow dynamics, we define two features:

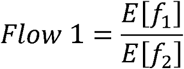

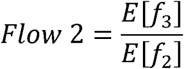

where *E*[.] denotes the expectation over time. Flow 1 quantifies the residual interactions between unresected EZ and non-EZ nodes, while Flow 2 captures the coupling between resected and preserved EZ regions. These measures provide interpretable, continuous metrics of how surgical resection impacts network connectivity relative to the predicted EZ.

### Predicting surgical outcomes using EZ centrality scores and network flow dynamics

To evaluate EZ centrality scores and network flow dynamics as interictal iEEG biomarkers of the EZ, we tested their efficacy in predicting surgical outcomes. We constructed the following three-dimensional feature vector:

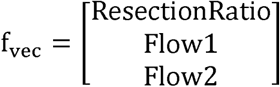

where

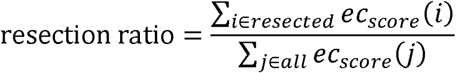

We trained and validated the surgical outcome prediction model using a multi-centre cohort of 163 patients with available postoperative outcome data (five patients were removed due to insufficient postoperative seizure outcomes follow-up). Because the cohort was imbalanced (approximately 70% seizure-free and 30% not seizure-free), model performance and generalization were evaluated using 10-fold stratified cross-validation. Patients were partitioned into 10 folds of roughly equal size, each preserving the class distribution, and models were trained on nine folds and evaluated on the held-out fold in each iteration. To avoid inflation of statistical significance due to within-patient dependence, all performance metrics were computed at the patient level. All analyses were performed using Python 3.14.

## Acknowledgements

The authors are deeply grateful to the patients who participated in this study and gave consent for the collection of their data for the advancement of knowledge and for a better future for epilepsy patients. We are indebted to Joyce H. Matsumoto, Lekha M. Rao, Rajsekar R. Rajaraman, Samuel Ahn, Maria Garcia Roca, Richard Le, Patrick Wilson, Cesar Dominguez, and Jimmy C Nguyen for their assistance with the study and sample acquisition.

## Data availability

The data and analysis code that support the findings of this study will be available upon publication. The EEG data are not publicly available due to privacy or ethical restrictions.

## References

1. Fiest KM, Sauro KM, Wiebe S, Patten SB, Kwon CS, Dykeman J, Pringsheim T, Lorenzetti DL, Jetté N. 2017. ‘Prevalence and incidence of epilepsy: A systematic review and meta-analysis of international studies’, Neurology, 88: 296–303. PMC5272794

2. Feigin VL, Vos T, Nichols E, Owolabi MO, Carroll WM, Dichgans M, Deuschl G, Parmar P, Brainin M, Murray C. 2020. ‘The global burden of neurological disorders: translating evidence into policy’, Lancet Neurol, 19: 255–265. PMC9945815

3. Kwan P, Brodie MJ. 2000. ‘Early identification of refractory epilepsy’, N Engl J Med, 342: 314–319.

4. Wiebe S, Blume WT, Girvin JP, Eliasziw M. 2001. ‘A randomized, controlled trial of surgery for temporal-lobe epilepsy’, N Engl J Med, 345: 311–318.

5. Zhang L, Hall M, Lam SK. 2023. ‘Comparison of long-term survival with continued medical therapy, vagus nerve stimulation, and cranial epilepsy surgery in paediatric patients with drug-resistant epilepsy in the USA: an observational cohort study’, Lancet Child Adolesc Health, 7: 455–462.

6. Rosenow F, Luders H. 2001. ‘Presurgical evaluation of epilepsy’, Brain, 124: 1683–1700.

7. Jayakar P, Gotman J, Harvey AS, Palmini A, Tassi L, Schomer D, Dubeau F, Bartolomei F, Yu A, Kršek P, Velis D, Kahane P. 2016. ‘Diagnostic utility of invasive EEG for epilepsy surgery: Indications, modalities, and techniques’, Epilepsia, 57: 1735–1747.

8. Gunnarsdottir KM, Li A, Smith RJ, Kang JY, Korzeniewska A, Crone NE, Rouse AG, Cheng JJ, Kinsman MJ, Landazuri P, Uysal U, Ulloa CM, Cameron N, Cajigas I, Jagid J, Kanner A, Elarjani T, Bicchi MM, Inati S, Zaghloul KA, Boerwinkle VL, Wyckoff S, Barot N, Gonzalez-Martinez J, Sarma SV. 2022. ‘Source-sink connectivity: a novel interictal EEG marker for seizure localization’, Brain, 145: 3901–3915. PMC10200292

9. Andrews JP, Gummadavelli A, Farooque P, Bonito J, Arencibia C, Blumenfeld H, Spencer DD. 2019. ‘Association of Seizure Spread With Surgical Failure in Epilepsy’, JAMA Neurol, 76: 462–469. PMC6459131

10. Widjaja E, Jain P, Demoe L, Guttmann A, Tomlinson G, Sander B. 2020. ‘Seizure outcome of pediatric epilepsy surgery: Systematic review and meta-analyses’, Neurology, 94: 311–321.

11. Kramer MA, Cash SS. 2012. ‘Epilepsy as a disorder of cortical network organization’, Neuroscientist, 18: 360–372. PMC3736575

12. Scott RC, Menendez de la Prida L, Mahoney JM, Kobow K, Sankar R, de Curtis M. 2018. ‘WONOEP APPRAISAL: The many facets of epilepsy networks’, Epilepsia, 59: 1475–1483.

13. Blumenfeld H, Varghese GI, Purcaro MJ, Motelow JE, Enev M, McNally KA, Levin AR, Hirsch LJ, Tikofsky R, Zubal IG, Paige AL, Spencer SS. 2009. ‘Cortical and subcortical networks in human secondarily generalized tonic-clonic seizures’, Brain, 132: 999–1012. PMC2724910

14. Burns SP, Santaniello S, Yaffe RB, Jouny CC, Crone NE, Bergey GK, Anderson WS, Sarma SV. 2014. ‘Network dynamics of the brain and influence of the epileptic seizure onset zone’, Proc Natl Acad Sci U S A, 111: E5321–5330. PMC4267355

15. Englot DJ, Hinkley LB, Kort NS, Imber BS, Mizuiri D, Honma SM, Findlay AM, Garrett C, Cheung PL, Mantle M, Tarapore PE, Knowlton RC, Chang EF, Kirsch HE, Nagarajan SS. 2015. ‘Global and regional functional connectivity maps of neural oscillations in focal epilepsy’, Brain, 138: 2249–2262. PMC4840946

16. Johnson GW, Doss DJ, Morgan VL, Paulo DL, Cai LY, Shless JS, Negi AS, Gummadavelli A, Kang H, Reddy SB, Naftel RP, Bick SK, Williams Roberson S, Dawant BM, Wallace MT, Englot DJ. 2023. ‘The Interictal Suppression Hypothesis in focal epilepsy: network-level supporting evidence’, Brain, 146: 2828–2845. PMC10316780

17. Bak P, Tang C, Wiesenfeld K. 1988. ‘Self-organized criticality’, Physical review A, 38: 364.

18. Nadeau RM, Foxall W, McEvilly TV. 1995. ‘Clustering and periodic recurrence of microearthquakes on the san andreas fault at parkfield, california’, Science, 267: 503–507.

19. Simkin M, Roychowdhury V. 2008. ‘A theory of web traffic’, Europhysics Letters, 82: 28006.

20. Simkin MV, Roychowdhury VP. 2011. ‘Re-inventing willis’, Physics Reports, 502: 1–35.

21. Farazmand M, Sapsis TP. 2019. ‘Extreme events: Mechanisms and prediction’, Applied Mechanics Reviews, 71: 050801.

22. Beggs JM, Plenz D. 2003. ‘Neuronal avalanches in neocortical circuits’, J Neurosci, 23: 11167–11177. PMC6741045

23. Stead M, Bower M, Brinkmann BH, Lee K, Marsh WR, Meyer FB, Litt B, Van Gompel J, Worrell GA. 2010. ‘Microseizures and the spatiotemporal scales of human partial epilepsy’, Brain, 133: 2789–2797. PMC2929333

24. Schevon CA, Weiss SA, McKhann G, Jr., Goodman RR, Yuste R, Emerson RG, Trevelyan AJ. 2012. ‘Evidence of an inhibitory restraint of seizure activity in humans’, Nat Commun, 3: 1060. PMC3658011

25. Wang SH, Siebenhühner F, Arnulfo G, Myrov V, Nobili L, Breakspear M, Palva S, Palva JM. 2023. ‘Critical-like Brain Dynamics in a Continuum from Second- to First-Order Phase Transition’, J Neurosci, 43: 7642–7656. PMC10634584

26. Simkin M, Roychowdhury V. 2010. ‘An explanation of the distribution of inter-seizure intervals’, Europhysics Letters, 91: 58005.

27. Wang S, Herzog ED, Kiss IZ, Schwartz WJ, Bloch G, Sebek M, Granados-Fuentes D, Wang L, Li JS. 2018. ‘Inferring dynamic topology for decoding spatiotemporal structures in complex heterogeneous networks’, Proc Natl Acad Sci U S A, 115: 9300–9305. PMC6140519

28. Bragin A, Engel J, Jr., Wilson CL, Fried I, Mathern GW. 1999. ‘Hippocampal and entorhinal cortex high-frequency oscillations (100--500 Hz) in human epileptic brain and in kainic acid--treated rats with chronic seizures’, Epilepsia, 40: 127–137.

29. Urrestarazu E, Chander R, Dubeau F, Gotman J. 2007. ‘Interictal high-frequency oscillations (100-500 Hz) in the intracerebral EEG of epileptic patients’, Brain, 130: 2354–2366.

30. Worrell GA, Parish L, Cranstoun SD, Jonas R, Baltuch G, Litt B. 2004. ‘High-frequency oscillations and seizure generation in neocortical epilepsy’, Brain, 127: 1496–1506.

31. Shi W, Shaw D, Walsh KG, Han X, Eden UT, Richardson RM, Gliske SV, Jacobs J, Brinkmann BH, Worrell GA, Stacey WC, Frauscher B, Thomas J, Kramer MA, Chu CJ. 2024. ‘Spike ripples localize the epileptogenic zone best: an international intracranial study’, Brain, 147: 2496–2506. PMC11224608

32. Jacobs J, Wu JY, Perucca P, Zelmann R, Mader M, Dubeau F, Mathern GW, Schulze-Bonhage A, Gotman J. 2018. ‘Removing high-frequency oscillations: A prospective multicenter study on seizure outcome’, Neurology, 91: e1040–e1052. PMC6140372

33. Roehri N, Pizzo F, Lagarde S, Lambert I, Nica A, McGonigal A, Giusiano B, Bartolomei F, Benar CG. 2018. ‘High-frequency oscillations are not better biomarkers of epileptogenic tissues than spikes’, Ann Neurol, 83: 84–97.

34. Zweiphenning W, Klooster MAV, van Klink NEC, Leijten FSS, Ferrier CH, Gebbink T, Huiskamp G, van Zandvoort MJE, van Schooneveld MMJ, Bourez M, Goemans S, Straumann S, van Rijen PC, Gosselaar PH, van Eijsden P, Otte WM, van Diessen E, Braun KPJ, Zijlmans M. 2022. ‘Intraoperative electrocorticography using high-frequency oscillations or spikes to tailor epilepsy surgery in the Netherlands (the HFO trial): a randomised, single-blind, adaptive non-inferiority trial’, Lancet Neurol, 21: 982–993. PMC9579052

35. Li A, Huynh C, Fitzgerald Z, Cajigas I, Brusko D, Jagid J, Claudio AO, Kanner AM, Hopp J, Chen S, Haagensen J, Johnson E, Anderson W, Crone N, Inati S, Zaghloul KA, Bulacio J, Gonzalez-Martinez J, Sarma SV. 2021. ‘Neural fragility as an EEG marker of the seizure onset zone’, Nature Neuroscience, 24: 1465–1474. PMC8547387

36. Jing J, Ge W, Hong S, Fernandes MB, Lin Z, Yang C, An S, Struck AF, Herlopian A, Karakis I, Halford JJ, Ng MC, Johnson EL, Appavu BL, Sarkis RA, Osman G, Kaplan PW, Dhakar MB, Arcot Jayagopal L, Sheikh Z, Taraschenko O, Schmitt S, Haider HA, Kim JA, Swisher CB, Gaspard N, Cervenka MC, Rodriguez Ruiz AA, Lee JW, Tabaeizadeh M, Gilmore EJ, Nordstrom K, Yoo JY, Holmes MG, Herman ST, Williams JA, Pathmanathan J, Nascimento FA, Fan Z, Nasiri S, Shafi MM, Cash SS, Hoch DB, Cole AJ, Rosenthal ES, Zafar SF, Sun J, Westover MB. 2023. ‘Development of Expert-Level Classification of Seizures and Rhythmic and Periodic Patterns During EEG Interpretation’, Neurology, 100: e1750–e1762. PMC10136013 Neurology.org/N for full disclosures.

37. Doss DJ, Shless JS, Bick SK, Makhoul GS, Negi AS, Bibro CE, Rashingkar R, Gummadavelli A, Chang C, Gallagher MJ, Naftel RP, Reddy SB, Williams Roberson S, Morgan VL, Johnson GW, Englot DJ. 2024. ‘The interictal suppression hypothesis is the dominant differentiator of seizure onset zones in focal epilepsy’, Brain, 147: 3009–3017. PMC11370787

38. Partamian H, Jahromi S, Corona L, Perry MS, Tamilia E, Madsen JR, Bolton J, Stone SSD, Pearl PL, Papadelis C. 2025. ‘Machine learning on interictal intracranial EEG predicts surgical outcome in drug resistant epilepsy’, NPJ Digit Med, 8: 138. PMC11880530

39. Tamilia E, Park EH, Percivati S, Bolton J, Taffoni F, Peters JM, Grant PE, Pearl PL, Madsen JR, Papadelis C. 2018. ‘Surgical resection of ripple onset predicts outcome in pediatric epilepsy’, Ann Neurol, 84: 331–346.

40. Matarrese MAG, Loppini A, Fabbri L, Tamilia E, Perry MS, Madsen JR, Bolton J, Stone SSD, Pearl PL, Filippi S, Papadelis C. 2023. ‘Spike propagation mapping reveals effective connectivity and predicts surgical outcome in epilepsy’, Brain, 146: 3898–3912. PMC10473571

41. Blumenfeld H. 2005. ‘Cellular and network mechanisms of spike-wave seizures’, Epilepsia, 46 Suppl 9: 21–33.

42. Khoshkhoo S, Vogt D, Sohal VS. 2017. ‘Dynamic, Cell-Type-Specific Roles for GABAergic Interneurons in a Mouse Model of Optogenetically Inducible Seizures’, Neuron, 93: 291–298. PMC5268075

43. Elahian B, Lado NE, Mankin E, Vangala S, Misra A, Moxon K, Fried I, Sharan A, Yeasin M, Staba R, Bragin A, Avoli M, Sperling MR, Engel J, Jr., Weiss SA. 2018. ‘Low-voltage fast seizures in humans begin with increased interneuron firing’, Ann Neurol, 84: 588–600. PMC6814155

44. Gliske SV, Irwin ZT, Chestek C, Hegeman GL, Brinkmann B, Sagher O, Garton HJL, Worrell GA, Stacey WC. 2018. ‘Variability in the location of high frequency oscillations during prolonged intracranial EEG recordings’, Nat Commun, 9: 2155. PMC5984620

45. von Ellenrieder N, Dubeau F, Gotman J, Frauscher B. 2017. ‘Physiological and pathological high-frequency oscillations have distinct sleep-homeostatic properties’, Neuroimage Clin, 14: 566–573. PMC5349616

46. Kini LG, Bernabei JM, Mikhail F, Hadar P, Shah P, Khambhati AN, Oechsel K, Archer R, Boccanfuso J, Conrad E, Shinohara RT, Stein JM, Das S, Kheder A, Lucas TH, Davis KA, Bassett DS, Litt B. 2019. ‘Virtual resection predicts surgical outcome for drug-resistant epilepsy’, Brain, 142: 3892–3905. PMC6885672

47. Jirsa V, Wang H, Triebkorn P, Hashemi M, Jha J, Gonzalez-Martinez J, Guye M, Makhalova J, Bartolomei F. 2023. ‘Personalised virtual brain models in epilepsy’, Lancet Neurol, 22: 443–454.

48. Zhang Y, Daida A, Liu L, Kuroda N, Ding Y, Oana S, Kanai S, Monsoor T, Duan C, Hussain SA, Qiao JX, Salamon N, Fallah A, Sim MS, Sankar R, Staba RJ, Engel J, Jr., Asano E, Roychowdhury V, Nariai H. 2025. ‘Self-supervised data-driven approach defines pathological high-frequency oscillations in epilepsy’, Epilepsia, 66: 4434–4450. PMC12266643

49. Zhang Y, Liu L, Ding Y, Chen X, Monsoor T, Daida A, Oana S, Hussain SA, Sankar R, Fallah A, Santana-Gomez C, Engel J, Staba RJ, Speier W, Zhang J, Nariai H, Roychowdhury V. 2024. ‘PyHFO: lightweight deep learning-powered end-to-end high-frequency oscillations analysis application’, J Neural Eng.

50. Ding Y, Zhang Y, Duan C, Daida A, Zhang Y, Kanai S, Lu M, Hussain SA, Staba RJ, Nariai H, Roychowdhury V. 2025. ‘PyHFO 2.0: an open-source platform for deep learning-based clinical high-frequency oscillations analysis’, J Neural Eng.

