## Extended Data Fig.1-7 for "Interictal Mini-Seizures: Recurrent Neuronal Synchronization Events Driven by the Epileptogenic Zone"

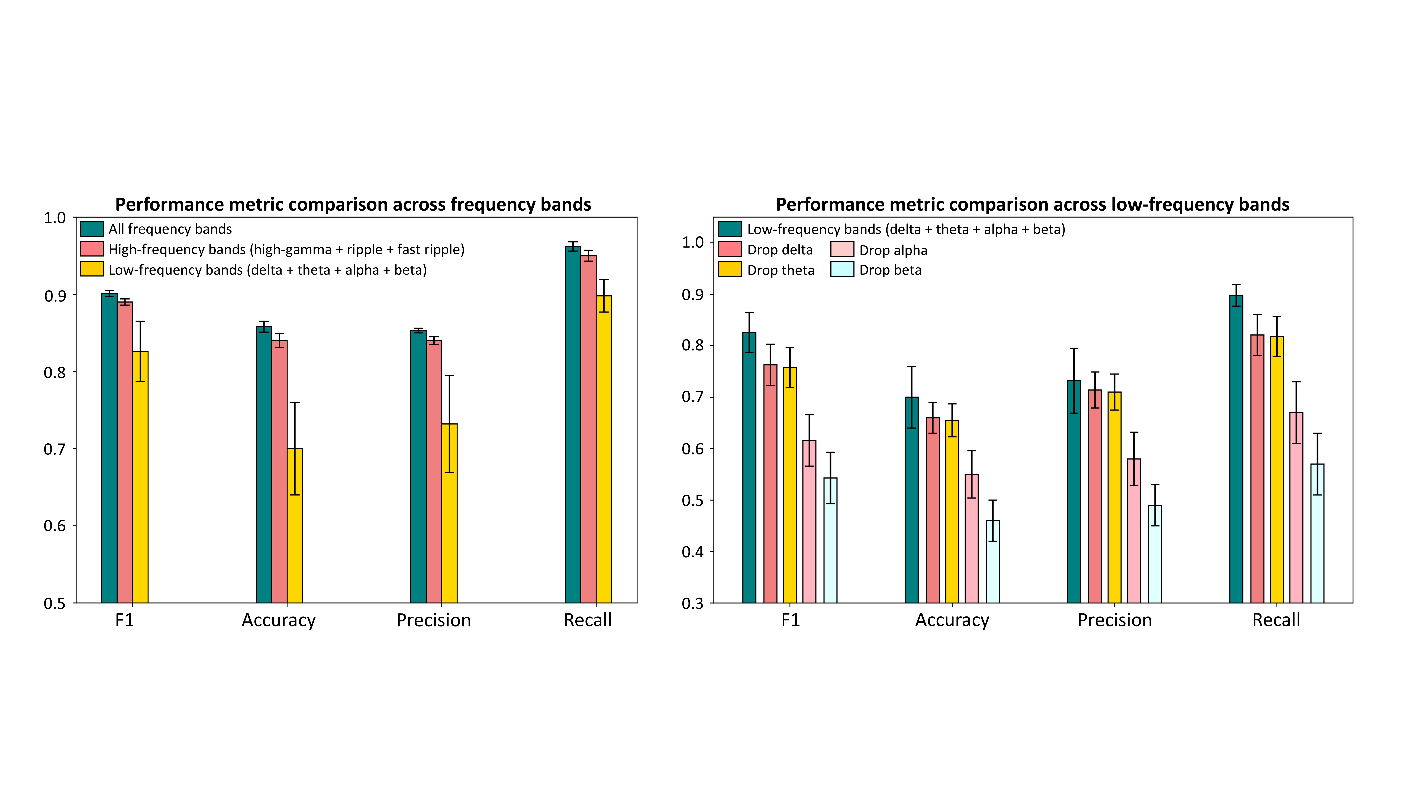

**Extended Data Figure 1. Frequency-band ablation reveals dominant contributions of high-frequency activity and critical roles of alpha and beta within low frequencies (n = 168 patients) in predicting surgical outcomes: elaborating on Fig 4(c).**

(A) Cross-validated performance comparison across frequency-band groupings for surgical outcome prediction (see Methods for outcome prediction model construction details). Models incorporating all frequency bands achieve the highest overall performance (F1 = 0.90, accuracy = 0.86, precision = 0.85, recall = 0.96). Restricting the model to high-frequency bands (high gamma, ripple, and fast ripple) preserves performance close to the full model across all metrics (e.g., F1 = 0.89; recall = 0.95), indicating that high-frequency activity carries most of the discriminative information. In contrast, models using low-frequency bands alone (delta, theta, alpha, and beta) show substantially reduced performance, most notably in accuracy (0.70) and precision (0.73), along with increased variability across validation folds. Recall that cross validation folds are across patients ensuring no patient gets to be on the training and test set on the same fold; thus, increased variability across folds correspond to increased inter-patient variability under low-frequency based prediction models. Together, these results suggest that low-frequency activity might be insufficient on its own.

(B) Ablation analysis within the low-frequency band set. Removing alpha results in marked reductions in performance (F1 from 0.83 to 0.62; recall from 0.90 to 0.67), while removing beta produces the largest decline across metrics (F1 = 0.54; accuracy = 0.46; recall = 0.57). In contrast, removal of delta or theta leads to only modest decreases in performance (F1 ≈ 0.76; accuracy ≈ 0.66), indicating that alpha- and beta-band activity contributes disproportionately to predictive power within the low-frequency range. Bars represent means across validation folds; error bars denote s.d. across validation folds.

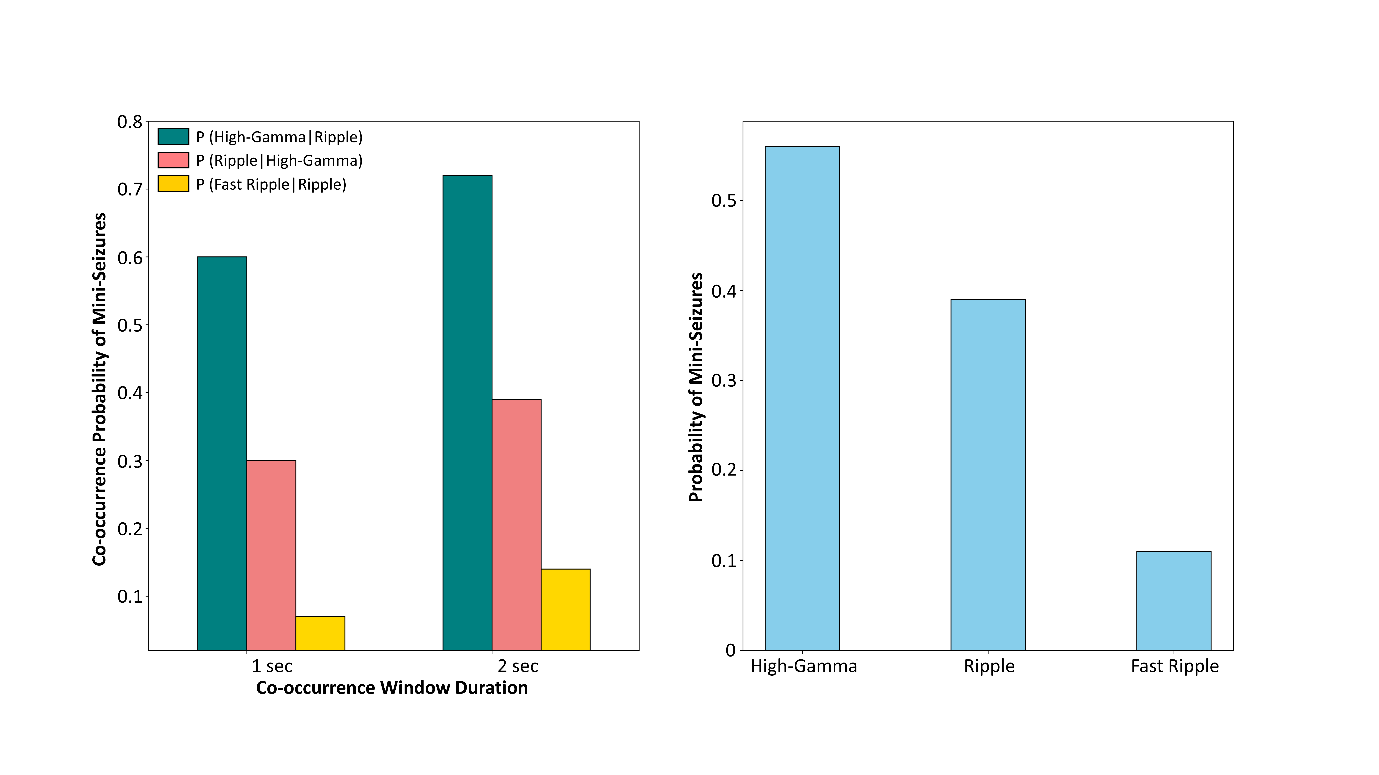

**Extended Data Figure 2. Mini-seizure activity is strongly coordinated across high-frequency bands, with ripple acting as a central co-occurrence hub (n = 168 patients): elaborating on Fig 1.**

(A) Conditional probabilities of mini-seizure co-occurrence across high-frequency bands for two temporal windows (1 s and 2 s), averaged across patients. Mini-seizure events detected (see Fig 1 and Methods for mini-seizure detection details) in one band are frequently accompanied by events in another, indicating robust cross-band coordination. Notably, relationships involving ripple are consistently strongest: the probabilities of observing mini-seizure in high-gamma or fast-ripple bands given mini-seizure observance in ripple band (P(hg | r) and P(fr | r)) remain high across both window durations. The stability of these patterns across 1 s and 2 s windows indicates that the observed coupling is not driven by a specific temporal parameter choice but reflects reproducible temporal coordination between bands.

(B) Band-wise occurrence probabilities of mini-seizures across high-frequency bands. All three bands—high gamma, ripple, and fast ripple—contribute substantially to mini-seizure detection. When considered together with the co-occurrence analysis in (A), these results indicate that ripple-associated activity provides the most consistent cross-band signature of mini-seizures, whereas high-gamma and fast-ripple events most often occur in conjunction with ripple rather than independently. Collectively, these findings demonstrate that mini-seizures are multiband high-frequency phenomena and identify ripple as a robust anchor for detecting coordinated high-frequency network activity across patients.

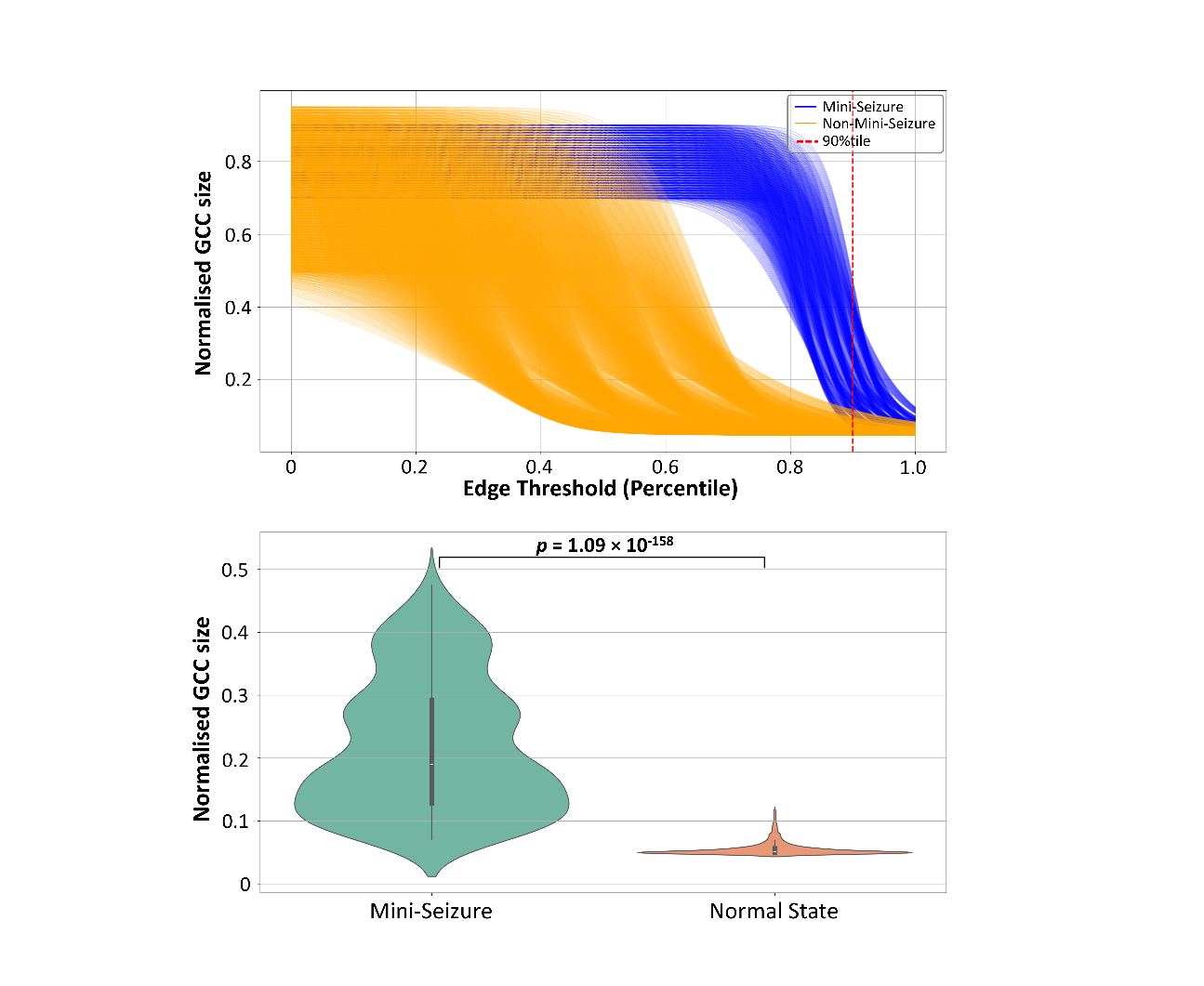

**Extended Data Figure 3. Selection of a 90th-percentile edge threshold yields a sparse yet connected network regime that maximizes separation between mini-seizure (MS) and non-MS events: elaborating on Fig 1(a) and (b).**

(Top) Across 168 patients, we used a 1-s sliding window to construct a sequence of synchronization networks and partitioned them into the two states: Normal state and mini-seizure state (see Methods for network construction and state identification details). Once we have the network sequences for the two states, then we track the network connectivity properties as a function of edge threshold (we drop all the network edges below the threshold) for each network state. (Left) Normalized giant connected component (GCC) size as a function of edge-weight percentile threshold. At low percentile thresholds, networks remain dense and near-saturated, with GCC size approaching 1 for both MS and non-MS events, limiting discriminability. As the threshold increases and weaker connections are progressively removed, networks enter a sparser regime in which group differences become pronounced. The 90th-percentile threshold (red dashed line) lies within this discriminative regime: MS events retain a substantially larger GCC than non-MS events at matched edge threshold percentile, indicating greater global network integration during MS. Thresholds above the 90th percentile approach an over-sparse regime in which GCCs begin to fragment into small components, increasing sensitivity to minor edge-weight fluctuations and reducing robustness, whereas lower thresholds retain many weak edges that obscure group differences.

(Bottom) Distribution of normalized GCC size at the 90th-percentile threshold. At this operating point, MS events show a marked upward shift relative to non-MS events, with an extremely significant between-group difference (two-sided t-test, P = 1.09 × 10⁻¹⁵⁸, as shown). Together, these results support the 90th-percentile threshold choice as a principled compromise that controls network density using a percentile-based rule, suppresses spurious weak connections while preserving the dominant connectivity backbone, and achieves large, statistically robust separation between MS and non-MS events without inducing network fragmentation.

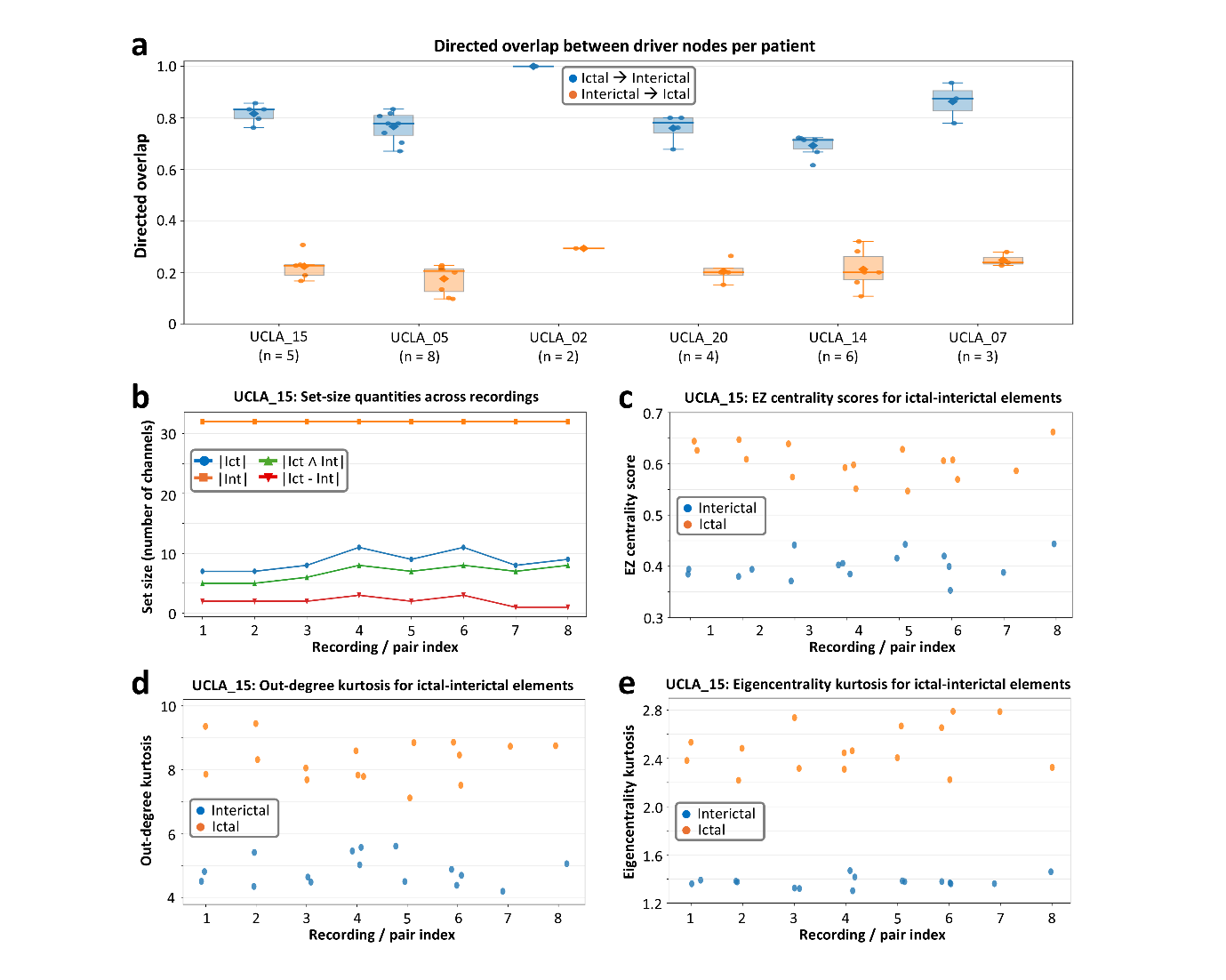

**Extended Data Figure. 4. Interictal driver nodes form an approximate superset of ictal driver nodes: elaborating on Fig. 3 (c, d)**

**(A)** Directed overlap between driver-node sets identified from ictal (Ict) and interictal (Int) recordings (see Extended Data Table 2 for ictal iEEG data), computed for each patient (P1–P6). The ictal→interictal overlap (blue), defined as $\mid Ict\cap Int\mid/\mid Ict\mid$, quantifies the fraction of ictal driver nodes contained within the interictal driver set. The interictal→ictal overlap (orange), defined as $\mid Ict\cap Int\mid/\mid Int\mid$, quantifies the fraction of interictal driver nodes contained within the ictal driver set. Distributions are computed across all pairwise ictal–interictal recording comparisons for each patient (sample size shown below each label). Box plots indicate the median and interquartile range, with whiskers extending to 1.5× the interquartile range; points denote individual recording pairs and diamonds indicate the mean. Across patients, consistently high ictal→interictal overlap combined with lower interictal→ictal overlap indicates that interictal driver sets retain most ictal drivers while additionally including extra channels, consistent with interictal drivers forming an approximate (near-)superset of ictal drivers at the recording-pair level.
**(B)** Example from patient P2 illustrating set-size relationships across eight ictal–interictal recording pairs, with a fixed interictal driver set size $\mid Int\mid$. For each pair, the sizes of $\mid Ict\mid$, $\mid Int\mid$, their intersection $\mid Ict\cap Int\mid$, and the ictal-only complement $\mid Ict-Int\mid=\mid Ict\mid-\mid Ict\cap Int\mid$are shown. Only a small subset of ictal drivers are absent from the interictal set, whereas the interictal set remains larger overall.
**(C)** EZ centrality scores for channels belonging to the ictal-only complement ($\mathrm{Ict}-\mathrm{Int}$) across the same P2 recording pairs. For each recording index, points represent individual channels. Interictal EZ centrality scores (blue) consistently occupy a lower range than ictal EZ centrality scores (orange), indicating systematic separation between interictal and ictal centrality estimates for this subset of channels.
**(D)** Out-degree kurtosis values for the same $\mathrm{Ict}-\mathrm{Int}$channels across P2 recordings. Interictal out-degree kurtosis (blue; approximately 4.2–5.7) is uniformly lower than ictal out-degree kurtosis (orange; approximately 7.1–9.9), consistent with stronger, more heavy-tailed outgoing connectivity dynamics during ictal epochs.
**(E)** Eigencentrality kurtosis values for $\mathrm{Ict}-\mathrm{Int}$channels across P2 recordings. Interictal eigencentrality kurtosis (blue; approximately 1.3–1.5) is lower than ictal eigencentrality kurtosis (orange; approximately 2.2–2.8) across all recordings, indicating increased peakedness and heavy-tailed centrality distributions during ictal periods.

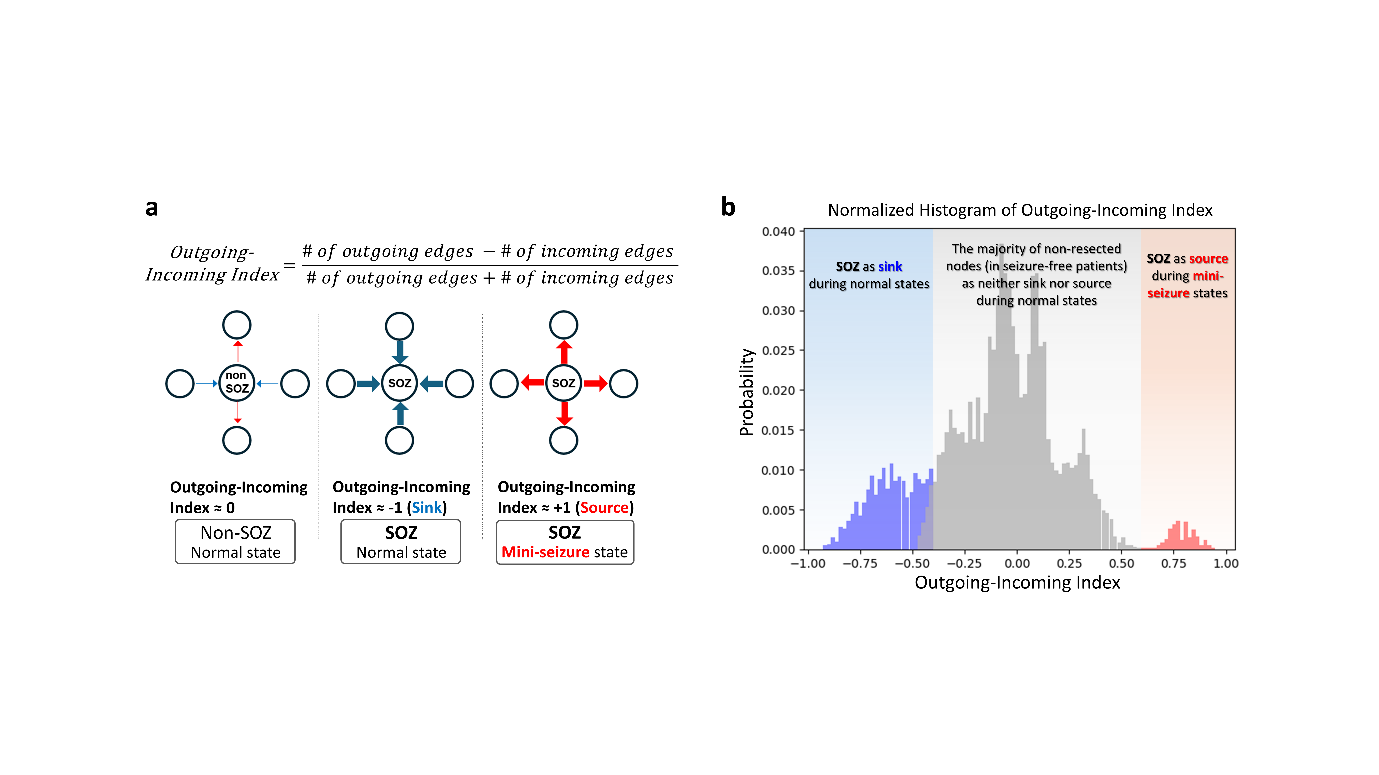

**Extended Data Figure. 5. The SOZ shifts from a sink state during normal interictal activity to a source state during mini-seizures: elaborating on Fig 3(a,b).**

(A) Definition of the Outgoing–Incoming index, computed from the balance of outgoing and incoming network connectivity for each node, as indicated by the formula. Positive values denote source-like behaviour (dominant outgoing influence), whereas negative values denote sink-like behaviour (dominant incoming influence).

(B) Probability distributions of the outgoing–incoming index across channels pooled from seizure free patients. During mini-seizures, SOZ channels are preferentially shifted toward source states, consistent with active driving of HSN dynamics. In contrast, during normal interictal states, SOZ channels predominantly occupy sink states, reflecting functional suppression. Preserved channels cluster near zero and do not exhibit a consistent source or sink bias.

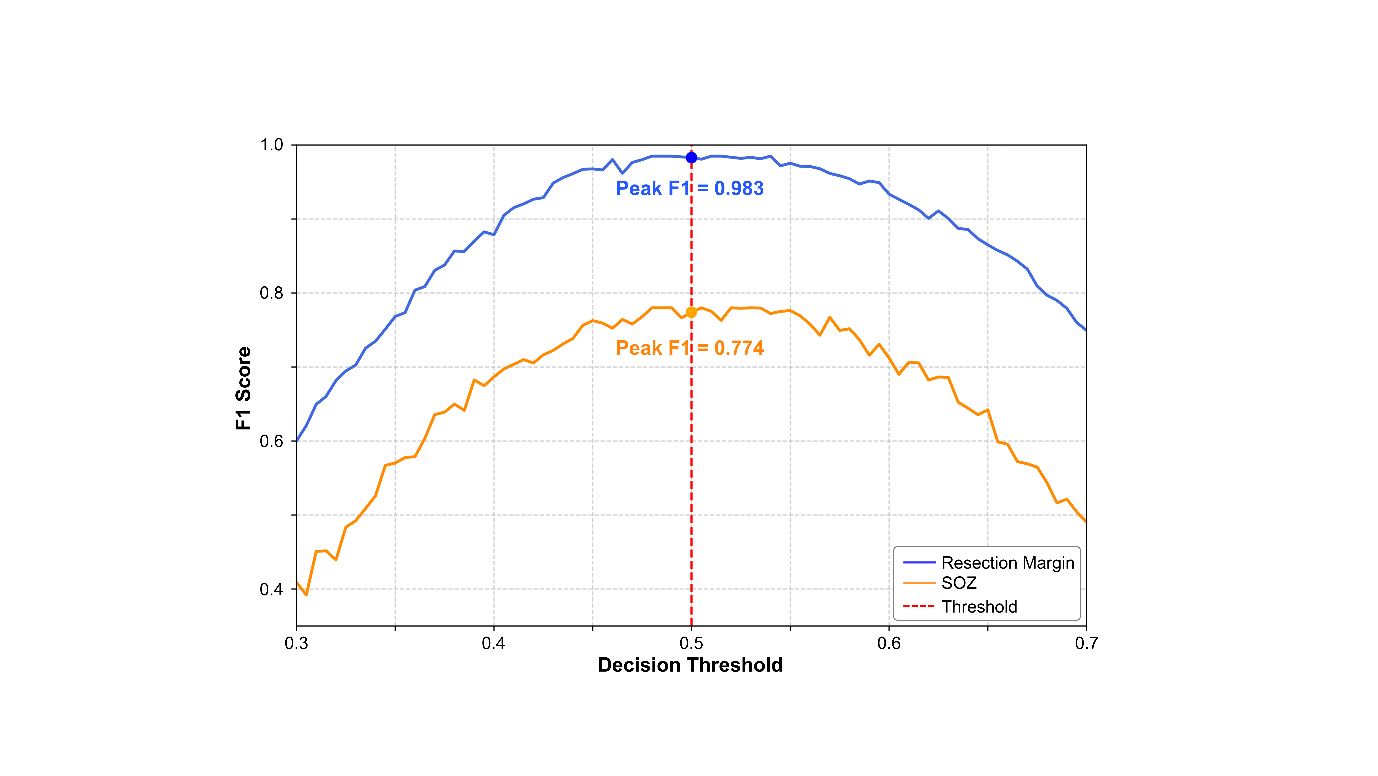

**Extended Data Fig. 6. Decision-threshold selection for resected channels and seizure onset zone (SOZ) prediction: elaborating on Fig 3(d).**

F1 score is shown as a function of the decision threshold for resected channels prediction (blue) and SOZ prediction (orange). A threshold of 0.5 (red dashed line) was selected as the operating point because it yields near-maximal F1 performance for both tasks simultaneously, achieving F1 ≈ 0.983 for resection-margin prediction and F1 ≈ 0.774 for SOZ prediction. Deviations from this threshold in either direction reduce F1 for both curves, consistent with the expected precision–recall trade-off. Notably, a threshold of 0.5 lies within a stable plateau around the peak of each curve, indicating robustness to small calibration or dataset shifts while preserving interpretability as the natural decision boundary for a calibrated probabilistic classifier.

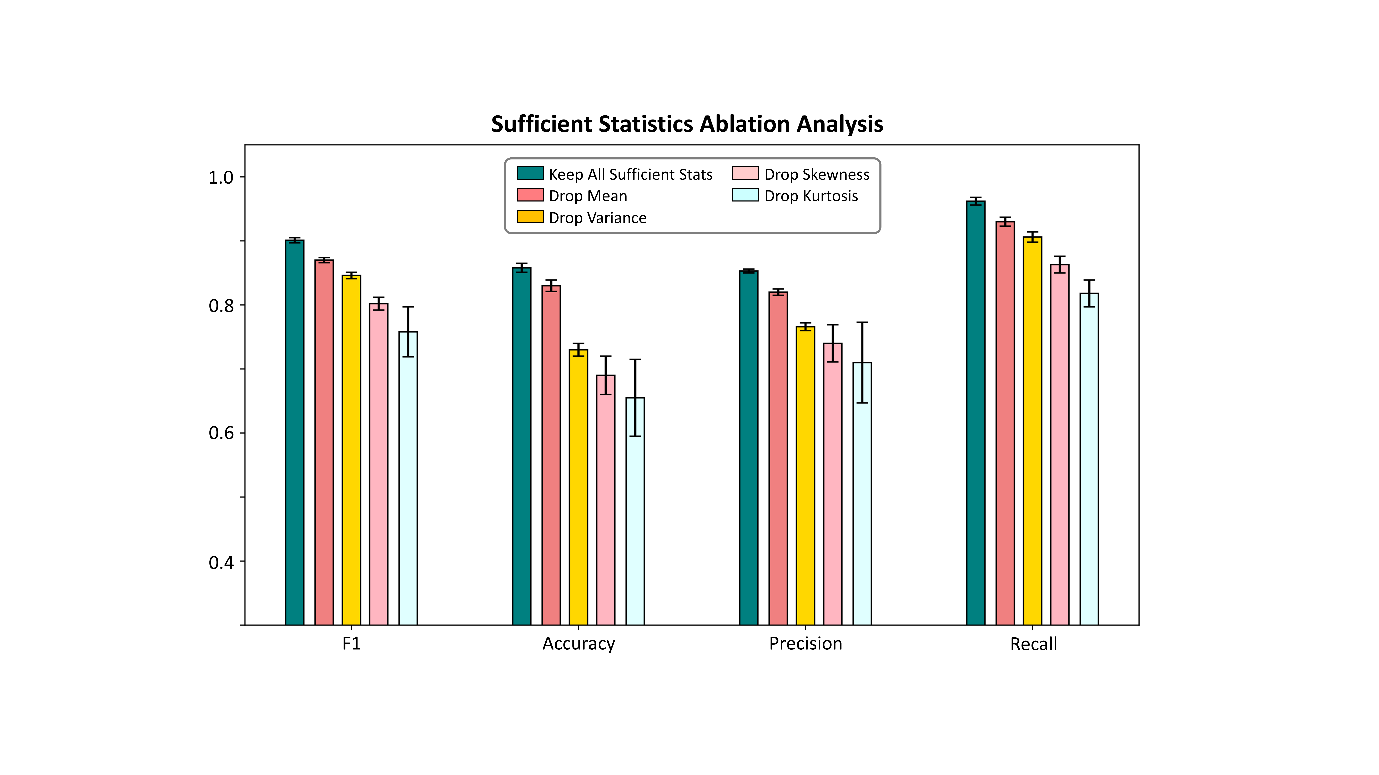

**Extended Data Figure. 7. Ablation of channel-level sufficient statistics identifies kurtosis as a critical determinant of EZ centrality–based surgical outcome prediction (n = 163 patients): elaborating on Fig 4(c)**

For each channel, time-varying network features—including out-degree, in-degree, scaled out-degree, scaled in-degree, and eigenvector centrality—are summarized over time using sufficient statistics (mean, variance, skewness, and kurtosis). These channel-level summaries are concatenated and used as input to an unsupervised class discovery procedure that yields an EZ centrality score for each channel. Patient-level features are then constructed from EZ centrality score ratios, together with additional network flow–based metrics, and used to train and evaluate the downstream surgical outcome prediction model (see Methods for details). Selective ablation of individual sufficient statistics prior to unsupervised discovery reveals that kurtosis is the most critical contributor. Removing kurtosis results in the largest outcome prediction performance degradation across metrics (F1 = 0.76, accuracy = 0.66, precision = 0.71, recall = 0.82) relative to the full model (F1 = 0.90, accuracy = 0.86, precision = 0.85, recall = 0.96), indicating that sensitivity to extreme temporal deviations and heavy-tailed dynamics in channel-level features is essential for stable EZ centrality estimation and accurate outcome prediction. Dropping skewness produces the next-largest decline in performance (F1 = 0.80, accuracy = 0.69, precision = 0.74, recall = 0.86), whereas removing variance (F1 = 0.85, accuracy = 0.73, precision = 0.77, recall = 0.91) or mean (F1 = 0.87, accuracy = 0.83, precision = 0.82, recall = 0.93) has a comparatively smaller effect. Bars represent mean performance across cross-validation folds; error bars denote variability across folds.
